# Model-based evaluation of the impact of a potential HIV cure on HIV transmission dynamics

**DOI:** 10.1101/2024.11.20.24317612

**Authors:** Alfredo De Bellis, Myrthe Willemsen, Giorgio Guzzetta, Ard van Sighem, Kim A.G.J. Romijnders, Peter Reiss, Maarten Schim van der Loeff, Janneke H.H.M. van de Wijgert, Monique Nijhuis, Mirjam Kretzschmar, Ganna Rozhnova

## Abstract

Biomedical research and clinical trials for curative HIV interventions are advancing rapidly. Understanding the potential impact of an HIV cure on transmission dynamics is key for its successful implementation at the population level. Using a mathematical model calibrated to the HIV epidemic among men who have sex with men (MSM) in the Netherlands as a case study, we tested the common hypothesis that an effective cure will contribute to HIV control and help end the epidemic. Guided by the target product profile (TPP) for an HIV cure, we evaluated the impact of two cure scenarios: (i) HIV remission, which entails suppression of the virus in the absence of ongoing antiretroviral treatment (ART) but carries a risk of viral rebound, and (ii) HIV eradication, which aims for complete removal of the virus. We found that either sustained HIV remission (with no risk of rebound) or HIV eradication could consistently reduce cumulative HIV incidence among MSM in the Netherlands by up to 50% for optimum TPP within eight years, relative to the baseline scenario without a cure. In contrast, transient HIV remission with a risk of rebound may drastically increase cumulative HIV incidence and disrupt the progress made toward HIV control under the current standard of care and prevention. These findings underscore the importance of the characteristics of curative HIV interventions in maximizing their benefits for public health and highlight the need to align HIV cure research with public health objectives to end the HIV epidemic.

## Introduction

Over the past decades, significant progress has been made toward achieving the United Nations Sustainable Development Goal of ending the HIV epidemic by 2030 [1]. Successes in reducing HIV transmission are particularly notable among men who have sex with men (MSM) in several Western European countries, including the Netherlands [2–4]. Since 2008, annual HIV diagnoses and HIV incidence among MSM in the Netherlands have declined by approximately 70% [2], largely due to public health interventions such as pre-exposure prophylaxis (PrEP) [5] and treatment as prevention [6].

Despite these advances, developing an HIV cure is a global health priority [7–9], as millions of people worldwide depend on lifelong antiretroviral treatment (ART) for viral suppression and a preventive vaccine is not yet available. People with HIV, including those in key populations such as MSM, experience impaired health-related quality of life despite effective ART [10–13]. Compared to individuals without HIV, people with HIV have worse physical and mental health due to chronic comorbidities and stigma [10, 12]. Emerging biomedical technologies, such as an HIV cure, have the potential to improve health and well-being of all people with HIV including MSM across various life domains, similar to how ART revolutionized HIV treatment in the past [7, 14, 15].

Biomedical research and human clinical trials for curative HIV interventions are advancing rapidly [16, 17]. To date, several patients have been cured of HIV through HIV-resistant stem cell transplants [18], but a scalable cure has yet to be developed. The consensus is that an acceptable and scalable cure for HIV will most likely require a combination of strategies targeting different aspects of HIV infection [7, 9, 19]. HIV eradication—aiming for the complete removal of HIV from the body—represents the ultimate goal for people with HIV [14, 20]. However, HIV remission, where the virus remains suppressed below transmissible levels without ongoing ART, could be a more attainable target for researchers. Strategies that are currently being developed may contribute to both HIV remission and HIV eradication [7, 19, 21–24].

The target product profile (TPP) for an HIV cure, a tool commonly used to guide drug development, has been devised by the International AIDS Society to align stakeholders on essential attributes for potential HIV cure interventions [9]. The TPP specifies key characteristics of a cure, including the target population, clinical efficacy, protection from re-infection, time to viral rebound, and monitoring requirements. According to the TPP’s minimum and optimum criteria, interventions resulting in either HIV remission or HIV eradication could be considered curative. Following the TPP framework, we refer to each of these cure scenarios as ‘a cure’ and to individuals who achieve it as ‘cured’.

The characteristics of a cure intervention could have major consequences not only for the well-being of people with HIV but also for virus transmission at the population level. It is often hypothesized that an effective cure could aid in controlling the HIV epidemic [7, 8, 24]. However, there is limited research investigating whether and how a cure would affect HIV transmission dynamics. For example, in an HIV remission scenario with a risk of viral rebound, transmission of the virus could occur. In an HIV eradication scenario, if a cure does not confer immunity, cured individuals may still be susceptible to re-infection. These considerations underscore the need for further investigation into the relationship between HIV cures and transmission dynamics.

From the early days of the HIV epidemic, mathematical modeling played an important role in advancing our understanding of HIV epidemiology, evaluating the impact of interventions, and shaping public health strategies [25–27]. For example, the seminal modeling study by Granich et al. [28] investigated the impact of test-and-treat on reducing HIV transmission, significantly influencing the international policy to end the HIV epidemic. Understanding how emerging HIV technologies, such as a potential cure, may affect the course of the epidemic is key to their successful implementation at the population level. Here, we considered the HIV epidemic among MSM in the Netherlands as a case study, where substantial progress has been made toward HIV control under the current standard of care and prevention. Using a mathematical model calibrated to epidemiological and sexual behavior data, we investigated the potential impact of curative interventions that would lead to either HIV remission or HIV eradication on the epidemic dynamics in this population. We explored under which assumptions regarding intervention characteristics these cure scenarios may increase or decrease HIV transmission among MSM in the Netherlands. Finally, we discussed the implications of our findings for the population-based implementation of cure interventions.

## Results

### Dynamics without cure

The model was fitted to and reproduced well the annual number of new HIV diagnoses and the estimated number of undiagnosed HIV infections among MSM from 2017 till 2022, reported by the Dutch HIV Monitoring Foundation (Stichting hiv monitoring, SHM) (Figure 1) [2]. The number of new HIV infections per 100,000 persons per year was estimated to decrease from 74 (95%CrI 33–127) in 2017 to 39 (95%CrI 14–67) in 2022. The estimated mean time to diagnosis was 28 months (95%CrI 24–33), consistent with the SHM data [2]. The model predicted that the proportions of diagnoses within 6 months, between 6 to 12 months, and more than 12 months since HIV infection were 22% (95%CrI 20%–24%), 14% (95%CrI 13%–16%), and 63% (95%CrI 60%–66%), respectively, also aligning with the SHM data [2]. The estimated HIV prevalence was almost constant, around 6%–7% throughout the considered period, which is in the range observed for MSM in Western Europe [29]. The model was additionally validated using independent data, not included in the model fitting, on the annual number of MSM on PrEP, the annual number of MSM on ART immigrating to the Netherlands from abroad, and the ART coverage among all MSM with HIV from 2017 till 2022 (Figure S1).

**Figure 1.**
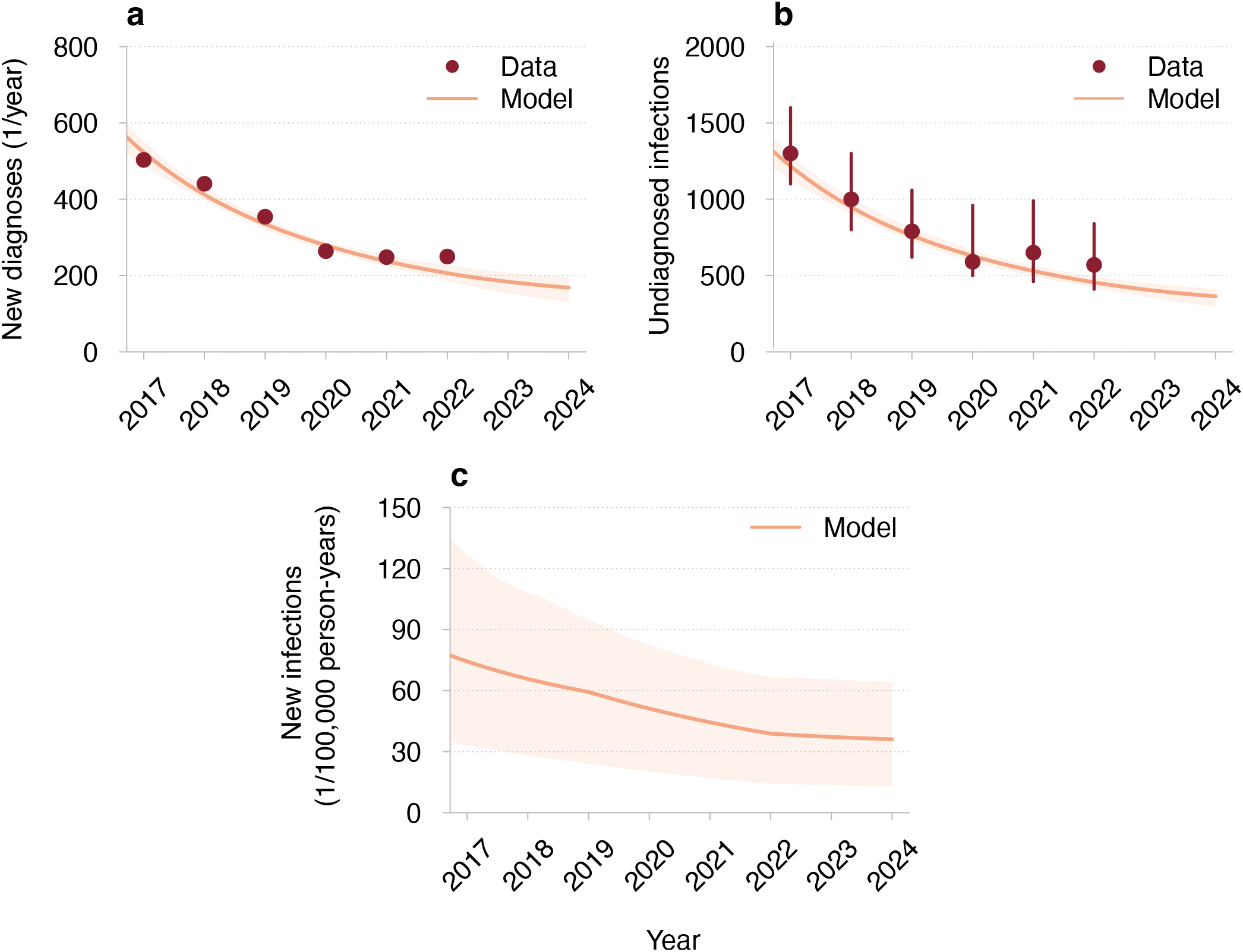
Model fit to HIV surveillance data for MSM in the Netherlands. (**a**) New HIV diagnoses, estimated number of undiagnosed HIV infections, and (**c**) new HIV infections. The red dots and the error bars correspond to the mean estimates and the 95% confidence intervals reported by SHM [2]. The mean trajectories estimated from the model are shown as orange lines. The shaded regions correspond to 95% credible intervals based on 100 samples from the joint posterior parameter distribution.

### Cure scenarios

Guided by the TPP [9], we developed a transmission model for the HIV remission and HIV eradication scenarios (see Figure 6 and Methods). The relation of these cure scenarios to the minimum and optimum TPP [9] is summarized in Table 1. In both scenarios, the cure was targeted at virologically suppressed individuals on ART (minimum TPP).

**Table 1.**
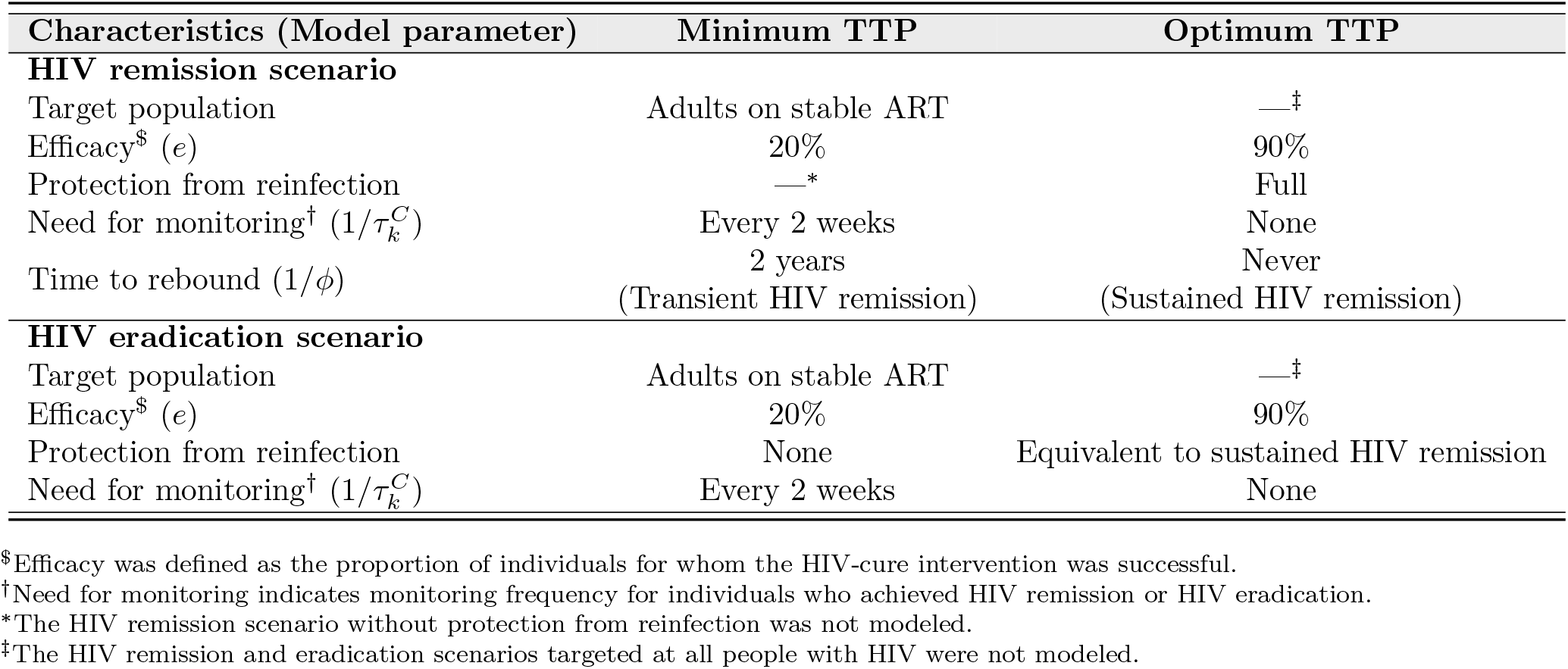
The relation of HIV cure scenarios to the minimum and optimum TPP for an HIV cure. Following the TPP framework [9], we refer to HIV remission and HIV eradication scenarios as ‘a cure’ and to individuals who achieve it as ‘cured’.

From an HIV dynamics perspective, the key difference between HIV remission and HIV eradication is that, in the remission scenario, the HIV reservoir within an individual is not completely removed. We defined ‘sustained’ HIV remission as a scenario where the virus remains below transmissible levels indefinitely (optimum TPP), whereas ‘transient’ HIV remission allows for possible viral rebound and potential onward transmission after a period of virus control of at least 2 years (minimum TPP). We assumed that individuals achieving HIV remission have full protection against re-infection (optimum TPP). In contrast, in the eradication scenario, all HIV including the intact and rebound-competent reservoir is fully removed. We further assumed that individuals achieving HIV eradication remain susceptible to re-infection (minimum TPP) and could start using PrEP if they have a high risk of HIV acquisition.

In both cure scenarios, we explored the entire range of acceptable values for cure characteristics outlined in the TPP, i.e., annual uptakes (proportion of eligible individuals receiving a cure intervention each year) of 10%, 50%, and 90%, and efficacy (proportion of individuals for whom the intervention was successful) of 20% (minimum TPP) and 90% (optimum TPP). Individuals who did not take up a cure intervention or for whom the intervention was unsuccessful remained on ART. Moreover, we considered three monitoring strategies for individuals who may experience a viral rebound after HIV remission or re-infection after HIV eradication: (i) no monitoring (optimum TPP), resulting in a mean (across all HIV stages) diagnostic delay of 28 months, which aligns with the estimated delay of HIV infections under the current standard of care [2]; (ii) PrEP-like monitoring, with a mean diagnostic delay of 3 months, similar to the testing interval for PrEP users [30]; and (iii) frequent monitoring (minimum TPP) every two weeks, as used in analytical treatment interruptions studies [31].

A cure intervention was assumed to be introduced in 2026, with maximum uptake reached within 3 years. We compared projections of HIV dynamics under the two cure scenarios and the no-cure scenario from 2026 to 2034. Changes in HIV incidence under the cure scenarios were always reported relative to the no-cure scenario. A detailed description of the model equations, parameters, and assumptions is provided in the Methods section, Figure 6, and the Supplementary Material.

### Dynamics for HIV remission scenario

We first explored how the dynamics under the HIV remission scenario might unfold based on different assumptions about the time to viral rebound. We simulated the model with mean rebound times of 2 years and 6 years, as well as the possibility of sustained HIV remission (i.e., no rebound). We assumed 90% intervention efficacy and 90% annual uptake, along with a 3-month diagnostic delay of rebounds. Projections of the epidemic dynamics for these parameters are shown in Figure 2. Sustained HIV remission, with no risk of rebound, resulted in the fewest new HIV infections compared to the no-cure scenario—8 (95%CrI 4–13) versus 33 (95%CrI 11–64) per 100,000 persons per year in 2034. If the time to viral rebound was 6 years, an estimated 23 (95%CrI 10–47) new HIV infections and 691 (95%CrI 664–715) new rebounds per 100,000 persons per year would occur in 2034. For a shorter rebound time of 2 years, new HIV infections and new rebounds increased to 41 (95%CrI 17–90) and 1,518 (95%CrI 1462–1570) per 100,000 persons per year in 2034. The HIV prevalence, including individuals in whom HIV remission was achieved but the viral reservoir was not fully removed, remained nearly constant regardless of the rebound time, while cure coverage (proportion of eligible individuals achieving HIV remission) was lower for shorter rebound times.

**Figure 2.**
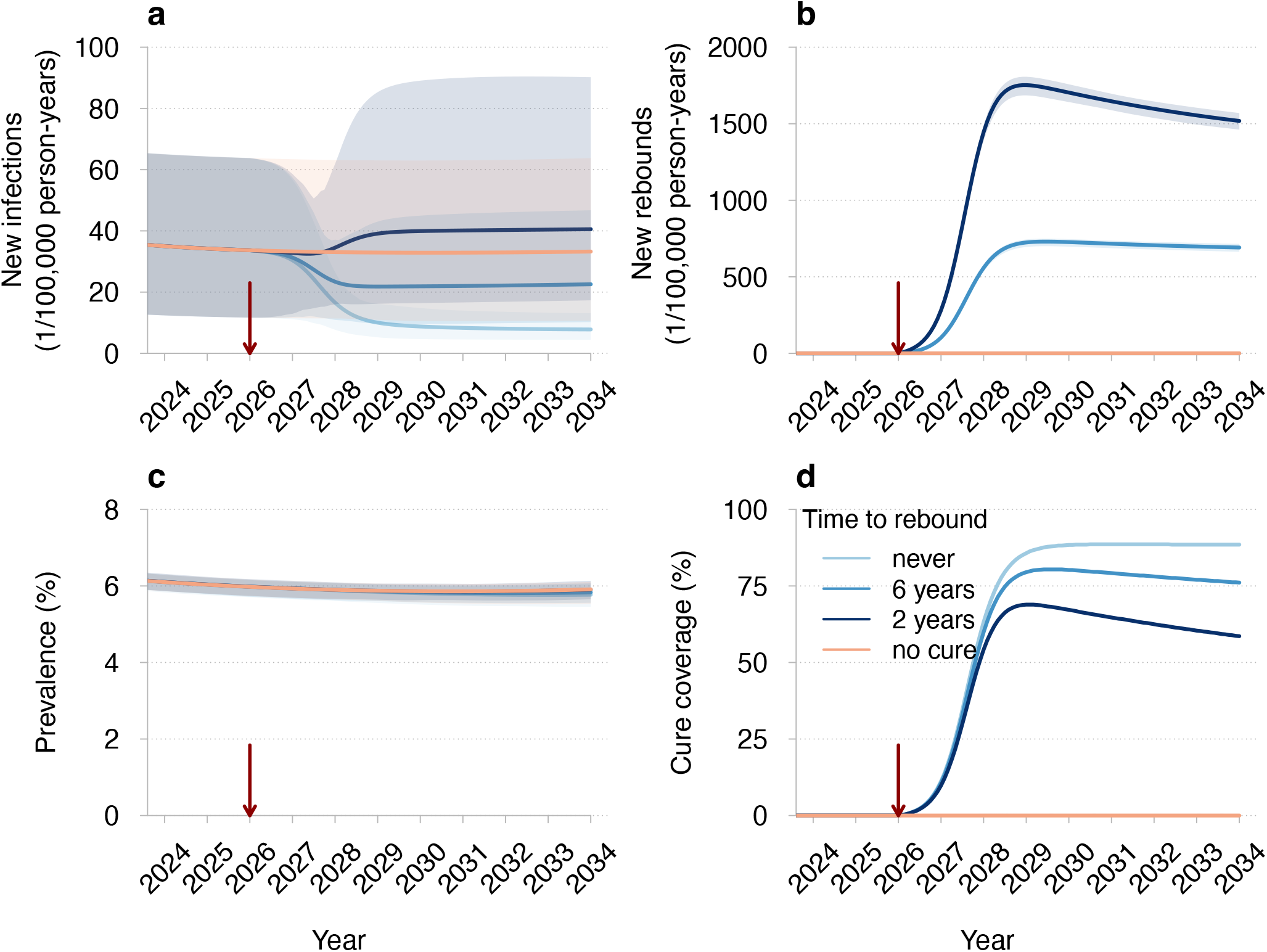
Projections of HIV dynamics under the HIV remission scenario. (**a**) New HIV infections, (**b**) new rebounds in individuals achieving HIV remission, (**c**) HIV prevalence (proportion of individuals with HIV), and (**d**) cure coverage (proportion of eligible individuals achieving HIV remission) for different times until viral rebound. The legend for different curves shown in (**d**) corresponds to all panels. The red vertical arrows indicate the cure introduction. The mean trajectories from the model are shown as solid lines. The shaded regions correspond to 95% credible intervals based on 100 samples from the joint posterior parameter distribution. Different shades of blue correspond to different times until viral rebound. The projections of the model without a cure are shown in orange. Parameters: efficacy of 90% (proportion of individuals for whom the intervention was successful), annual uptake of 90% (proportion of eligible individuals receiving the intervention each year), and a 3-month diagnostic delay of rebounds in individuals who achieved HIV remission.

### Impact of HIV remission under varied intervention characteristics

We further systematically compared the impact of HIV remission under varied efficacy, uptake, time to rebound, and diagnostic delay from its introduction in 2026 to 2034 (Figure 3). Sustained HIV remission consistently resulted in fewer HIV infections, regardless of diagnostic delay (Figure 3, top rows). Mean reductions in cumulative HIV infections over this period, compared to the no-cure scenario, ranged from 52% for 90% uptake and 90% efficacy to 3% for 10% uptake and 20% efficacy. Intermediate reductions in HIV infections were predicted for all other parameter combinations.

**Figure 3.**
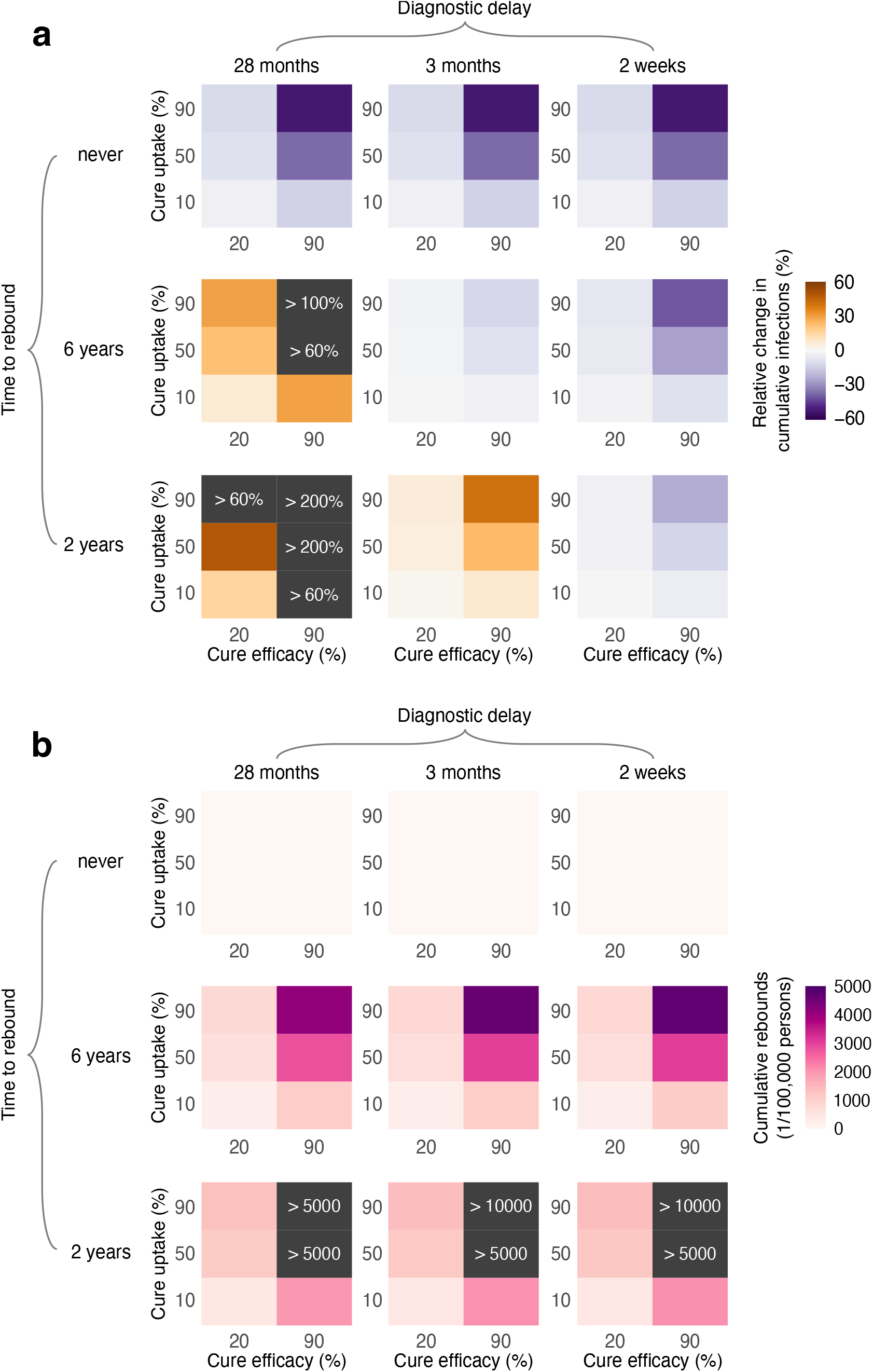
Impact of HIV remission on HIV dynamics under varied intervention characteristics. (**a**) Mean change in cumulative HIV infections relative to the no-cure scenario and (**b**) mean cumulative rebounds from the introduction of HIV remission scenario in 2026 to the end of the simulation in 2034.

Whether transient HIV remission increased or decreased HIV infections from 2026 to 2034, compared to the no-cure scenario, depended on diagnostic delay and rebound time (Figure 3, middle and bottom rows). In the absence of a specific monitoring strategy to diagnose rebounds (i.e., a 28-month diagnostic delay of HIV infections observed under the current standard of care), the introduction of this intervention consistently led to more HIV infections. Larger increases in infections were predicted for shorter rebound time, and higher uptake and efficacy. Conversely, with frequent monitoring of individuals who achieved HIV remission (i.e., a 2-week diagnostic delay of rebounds), cumulative HIV infections were consistently reduced compared to the no-cure scenario. The largest mean reductions in cumulative HIV infections were between 41% for a rebound time of 6 years and 23% for a rebound time of 2 years. However, an estimated 4,695 (95% CrI 4,485–4,847) and 11,310 (95% CrI 10,836–11,653) rebounds per 100,000 persons were predicted to occur alongside these maximum reductions in HIV infections. During the same period, only 561 (95% CrI 184–1,074) HIV infections per 100,000 persons were estimated to occur in the no-cure scenario.

### Dynamics for HIV eradication scenario

Similar to HIV remission, we first assessed transmission dynamics under the HIV eradication scenario, considering 90% efficacy and a 3-month diagnostic delay for re-infections. The introduction of HIV eradication led to a consistent reduction in the number of new HIV infections in naive individuals, with values varying based on cure uptake (Figure 4). For 90% uptake, the estimated number of new HIV infections decreased to 8 (95% CrI 4–13) per 100,000 persons per year in 2034, which is comparable to the outcome of sustained HIV remission. For lower uptakes, new HIV infections also declined, albeit less markedly, reaching 9 (95% CrI 5–14) and 22 (95% CrI 9–39) per 100,000 persons per year for 50% and 10% uptake, respectively. Notably, across the entire range of uptakes considered, the estimated number of re-infections in cured individuals remained low, approaching only about 2 re-infections per 100,000 persons per year in 2034. Furthermore, unlike in the HIV remission scenario, HIV prevalence markedly dropped after the introduction of HIV eradication, reaching 3.60% (95%CrI 3.38%–3.74%) for 10% uptake and falling to less than 1% for uptakes above 50%. The model also indicated that for lower uptakes, equilibrium in HIV dynamics had not yet been reached, with new HIV infections projected to decline further after 2034 (e.g., falling below 22 per 100,000 persons for 10% uptake).

**Figure 4.**
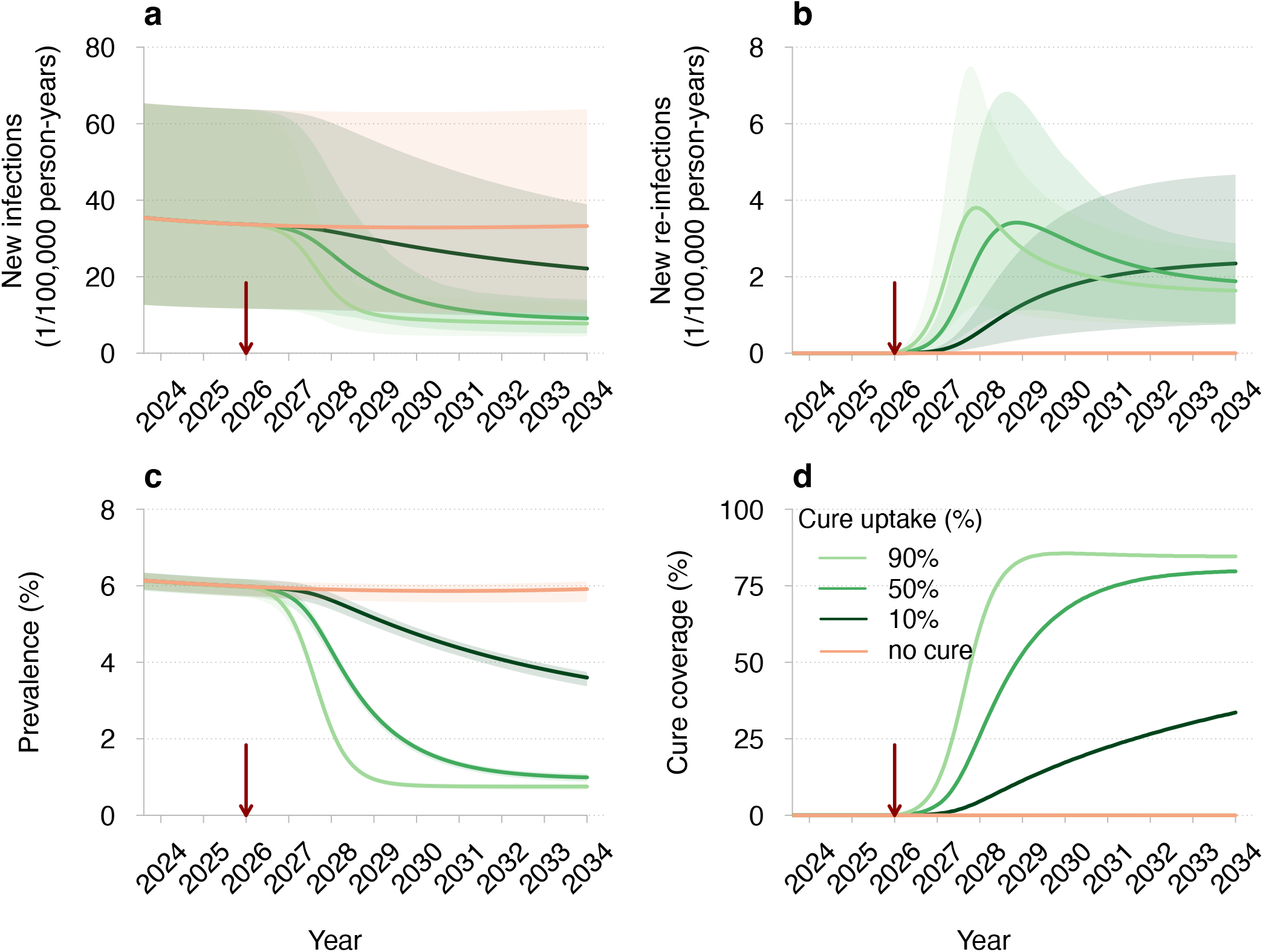
Projections of HIV dynamics under the HIV eradication scenario. (**a**) New HIV infections (primary infections in naive individuals), (**b**) new HIV re-infections (secondary infections in cured individuals), HIV prevalence (proportion of individuals with HIV), and (**d**) cure coverage (proportion of cured individuals among all eligible) for different cure uptakes. The legend for different curves shown in (**d**) corresponds to all panels. The red vertical arrows indicate the cure introduction. The mean trajectories from the model are shown as solid lines. The shaded regions correspond to 95% credible intervals based on 100 samples from the joint posterior parameter distribution. Different shades of green correspond to different cure uptakes. The projections of the model without a cure are shown in orange. Parameters: efficacy of 90% (proportion of individuals for whom the intervention was successful) and a 3-month diagnostic delay of re-infections in individuals who achieved HIV eradication.

### Impact of HIV eradication under varied intervention characteristics

A systematic comparison of the impact of HIV eradication under varied efficacy, uptake, and diagnostic delay of re-infections is shown in Figure 5. Similar to sustained HIV remission, HIV eradication consistently reduced HIV infections in naive individuals, regardless of diagnostic delay. Like before, the largest and smallest reductions in cumulative HIV infections over the 2026-2034 period were observed at the extreme values of uptakes and efficacy, with the reduction ranging from 3% to 52% for all parameter combinations explored. For all diagnostic delays, the estimated number of cumulative re-infections remained low, not exceeding 16 per 100,000 persons over 8 years.

**Figure 5.**
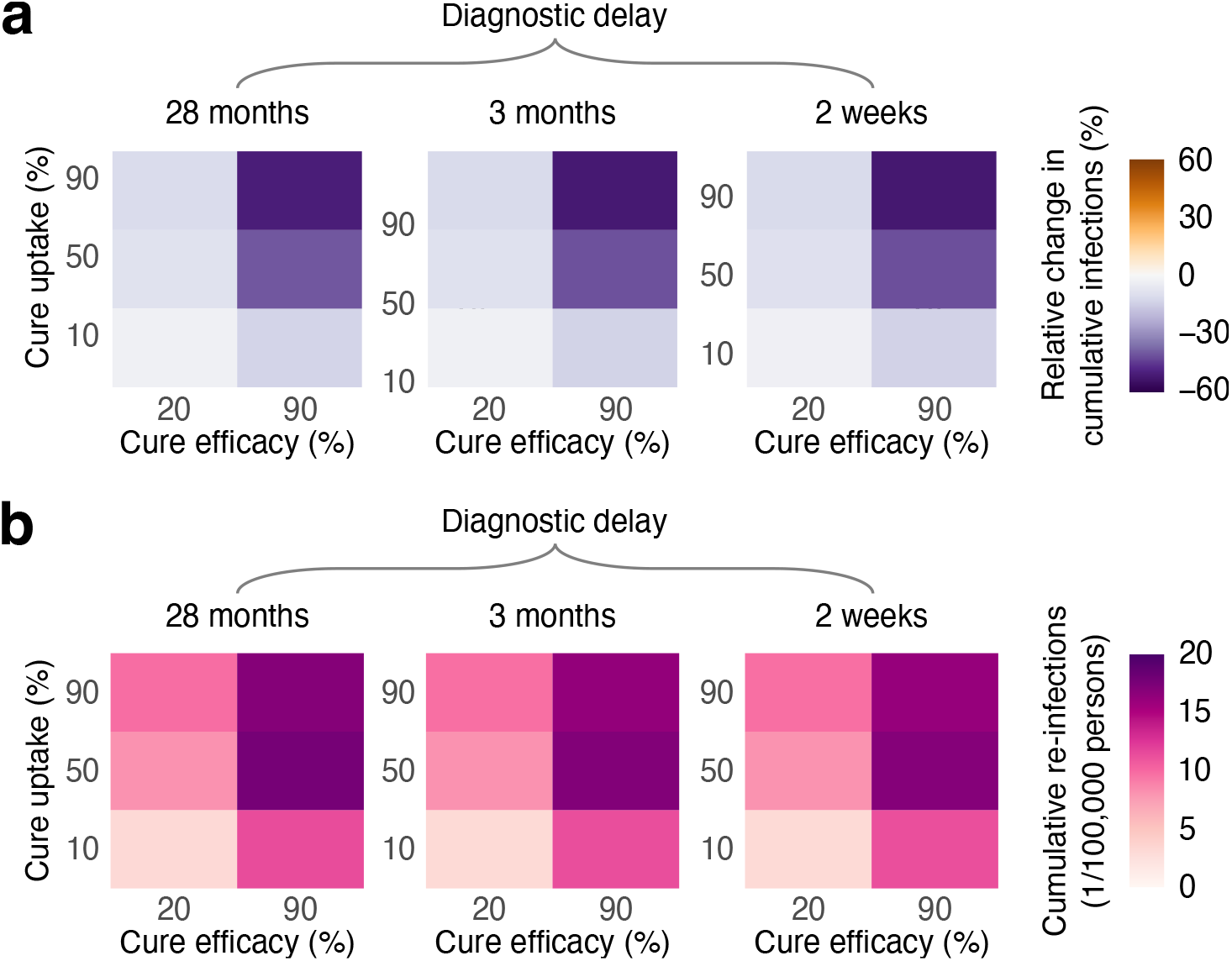
Impact of the HIV eradication on HIV dynamics under varied cure characteristics. (**a**) Mean change in cumulative HIV infections (primary infections in naive individuals) relative to the no-cure scenario and (**b**) mean cumulative HIV re-infections (secondary infections in cured individuals) from the introduction of HIV eradication scenario in 2026 to the end of the simulation in 2034. The color bar scale is the same as that in Figure 3 for direct comparison.

### Robustness and sensitivity analyses

Our results remained robust across different years of cure introduction. A potential increase in sexual risk behavior among MSM in response to a cure based on survey data did not qualitatively affect the overall outcomes. Specifically, eight years post-cure, the number of new HIV infections was always lower for HIV eradication compared to the no-cure scenario, but this was not the case for transient HIV remission. Sensitivity analyses regarding the infectivity of individuals after viral rebound indicated that our projections for HIV remission were the most optimistic. In contrast, the results for HIV eradication were consistent across varying assumptions about the infectivity of re-infections. The qualitative insights into the impact of cure scenarios relative to the no-cure scenario were also robust to potential changes in the underlying trend in HIV incidence, particularly when new HIV infections started to increase from 2022 onward. Further details of these and additional sensitivity analyses are given in the Supplementary Material.

## Discussion

In light of rapidly advancing HIV cure research [19], anticipating the potential population-level impact of cure implementation is essential before effective interventions become available. From a public health perspective, an effective cure should align with the United Nations Sustainable Development Goal of ending the HIV epidemic [1]. Indeed, there is a prevailing assumption that a cure will contribute to epidemic control [7, 8, 24]. In Western European countries where HIV epidemics are concentrated among MSM and HIV incidence is already low, this assumption implies that the implementation of a cure should not disrupt the progress made in HIV control under the current prevention and treatment standards.

We found that introducing either cure scenario could alter the course of the HIV epidemic among MSM in the Netherlands. The impact differed between sustained HIV remission or HIV eradication, which both have the potential to further reduce HIV transmission, and transient HIV remission, which could increase HIV transmission. This divergence in the potential outcomes of cure scenarios underscores the importance of evaluating their public health impacts, which, for now, can only be done through mathematical modeling [32]. Specifically, the model projections indicated that while sustained HIV remission consistently resulted in a decrease in cumulative HIV incidence over eight years (by up to 50% for optimum TPP), transient HIV remission could drastically increase new infections. Notably, the increase in incidence was observed for plausible parameter combinations that might be achievable in a real-world setting (e.g., a 3-month diagnostic delay, which matches the testing interval for PrEP users [30], and the TPP’s minimum requirement of 2 years before relapse [9]). HIV incidence under transient HIV remission increased in the model because individuals who experienced viral rebound could transmit the virus before being diagnosed. This led to a counterintuitive effect where higher incidence was observed for higher cure uptakes and efficacies. The more individuals achieved HIV remission, the more rebounds occurred, leading to an increase in new infections. This effect could be mitigated by developing HIV remission strategies with longer times to rebound or through stricter monitoring. However, even in scenarios where transient HIV remission reduces HIV incidence, managing numerous rebound cases may pose a challenge in the real world and lead to adverse health outcomes for patients. People with HIV in the Netherlands and Australia have also raised concerns about transient HIV remission because of fear of transmitting the virus to their partners and stigma [14, 20]. Frequent rebound episodes would require ongoing efforts for rapid viral load monitoring and timely diagnosis of rebounds to maximize the public health benefits of any HIV remission strategy. The development of a simple point-of-care or at-home diagnostic for detecting rebounds may be a potential solution mentioned by experts who developed the TPP for an HIV cure [9]. In contrast, the HIV eradication scenario presents a more optimistic outlook for HIV dynamics. Our findings suggest that this scenario would always reduce HIV incidence among naive individuals, with projected cumulative new infections decreasing similarly to the sustained HIV remission scenario (by up to 50% for optimum TPP) within eight years. Importantly, the HIV eradication scenario would maintain a low number of re-infections, which is relevant for those MSM who are particularly vulnerable due to their sexual risk behavior [33]. The main reason for few re-infections under the HIV eradication scenario and a potentially large number of rebounds under the transient HIV remission scenario is that the rate of re-infections depends on the population with HIV, while rebounds occur at a constant rate among individuals who achieved HIV remission.

It is recognized that mathematical modeling can help advance HIV cure research [32]. However, few studies have modeled the impact of cure strategies at the population level. To our knowledge, this is the first study to assess the potential impact of an HIV cure on an epidemic among MSM in a Western European country with low HIV incidence. We ensured the reliability of our model to project HIV dynamics without a cure by inferring key model parameters and conducting model validation on multiple datasets. Unlike other studies assessing interventions for HIV control in low-incidence settings [34–36], our model accounted for the openness of the MSM population by incorporating immigration of individuals to the Netherlands who acquired HIV abroad. The strength of our modeling approach lies in its ability to project HIV dynamics without requiring precise information on the biological mechanisms underlying a cure. Instead, our analyses were guided by the TPP, using the full range of acceptable values for several cure characteristics [9].

There is concern that the introduction of a cure could shift perceptions of HIV risk, severity, and prevention, as observed when ART became widely available [37]. Our study is the first to incorporate actual survey data on potential behavioral changes among MSM following the introduction of a cure into sensitivity analyses. Our findings suggest that risk compensation could further reduce the effectiveness of the transient HIV remission scenario while having minimal impact on the outcomes of the HIV eradication scenario. Therefore, real-world cure interventions may need to be accompanied by additional prevention strategies to address the potential change in sexual behavior. While our findings are particularly relevant for similar HIV epidemics among MSM in Western European countries, where HIV incidence is low, they align qualitatively with two previous modeling studies for a generalized epidemic in heterosexual populations in Africa [38, 39]. These studies, though not formally fitted to data, also suggested that a remission-like intervention without sustained viral suppression could lead to an increase in HIV incidence. However, our results differ from [39], as we found that the timing of cure introduction did not alter the outcomes of cure scenarios. This discrepancy may be explained by the fact that the HIV epidemic in the Netherlands was estimated to be close to reaching a low stable level of incidence, unlike the more dynamic epidemic in South Africa. Our study has several limitations. First, we used a classical deterministic compartmental model, which assumed an exponential distribution for the time until viral rebound. Although this distribution may not perfectly capture relapse timing, it provides a way to compare cure scenarios conceptually, given the limited empirical data available. While a stochastic model might yield different quantitative outcomes, our primary conclusions would likely remain robust. Additionally, our projections are not intended as precise forecasts, as future changes in the national PrEP program, sexual behavior or migration trends could influence the quantitative predictions for the no-cure scenario, potentially leading to an increase in HIV incidence. Nevertheless, our sensitivity analyses showed that the qualitative insights regarding the impact of cure scenarios relative to the no-cure scenario are expected to hold despite such changes. Second, despite the formulation of the TPP, uncertainty remains around several biological parameters for cured individuals. Based on data from ART interruption studies, we assumed that the infectivity of individuals following viral rebound would be similar to that observed during the chronic stage of HIV infection [40, 41]. Conversely, we assumed the infectivity of individuals re-infected after HIV eradication would resemble the acute stage of HIV infection, similar to what occurs with superinfection by a different HIV subtype [42] or reinfection with hepatitis C virus [33,43]. While these assumptions are biologically plausible, they remain hypothetical. Our sensitivity analyses demonstrated that high infectivity could undermine the effectiveness of the transient HIV remission scenario and should be carefully considered when developing a cure. Lastly, in line with the call for equitable HIV cure solutions [44–47], our analysis did not focus on targeting cure strategies to specific population groups based on behavior or HIV status. Instead, in agreement with the inclusion criteria for many HIV cure trials [48] and the minimum requirement in the TPP [9], our model assumes that a cure is administered to individuals on ART. Since most MSM with HIV in the Netherlands are diagnosed and receive effective treatment promptly upon diagnosis [2, 49], this assumption likely has minimal impact on our findings.

In summary, our study suggests that both HIV eradication and sustained HIV remission have the potential to reduce new HIV infections, contributing to the United Nations Sustainable Development Goal of ending the HIV epidemic, while transient HIV remission could increase infections if rebounds are not promptly monitored. These findings emphasize the need for further research to ensure strategic development and effective implementation of cure interventions.

## Methods

### Overview

A classical deterministic compartmental model was calibrated using sexual behavior and HIV surveillance data for MSM in the Netherlands. The model was validated against independent data not used in the fitting process. This calibrated model was then applied to explore the potential impact of an HIV cure on epidemic dynamics at the population level.

### Data

#### Sexual behavior data

We used sexual behavior data from a cross-sectional survey conducted from October 2021 to June 2022 [50]. The survey aimed to assess the anticipated impact of an HIV cure on quality of life, sexual satisfaction, stigma, and sexual and preventive behaviors among people with HIV and key populations in the Netherlands. From this survey, we extracted and analyzed responses from *n* = 529 MSM participants who provided information on the number of sexual partners in the last six months and condom use, and hypothetical changes in these variables following the introduction of HIV remission and HIV eradication scenarios. Additional details on the use of behavioral data are provided in the Model Calibration section and Supplementary Material.

#### HIV surveillance data

In the Netherlands, care for people with HIV is provided by 24 designated HIV treatment centers. The Dutch HIV Monitoring Foundation (Stichting hiv monitoring, SHM) was appointed by the Ministry of Health, Welfare and Sport to continuously monitor the HIV epidemic in the Netherlands and report on all aspects of HIV care. Since 1998, SHM has collected data in the ATHENA cohort from over 98% of all people with HIV who are receiving care in the Netherlands [51]. SHM annually reports on the number of new HIV diagnoses and publishes estimates of the number of newly acquired HIV infections and the undiagnosed population. We used the SHM data for MSM from 2015 to 2022 on the annual number of new HIV diagnoses, the annual number of individuals on ART immigrating to the Netherlands from abroad, and ART coverage [2]. We also used the SHM estimates of undiagnosed HIV infections, calculated using the European Centre for Disease Prevention and Control HIV Platform tool [2, 52]. Data on the number of PrEP users in the national PrEP program from the national sexually transmitted infections surveillance database [53], available from June 2019 to April 2022, were taken from [34].

### Transmission model

#### Model without cure

The model described the sexual transmission of HIV among MSM in the Netherlands. The population was stratified into four risk groups based on the average number of new sexual partners per year, *c*_*l*_ [35]. We denoted risk groups with subscript *l* = 1, 2, 3, 4, where *l* = 1 represented the group with the lowest number of partners and *l* = 4 with the highest. Individuals did not change risk groups. The population in risk group *l* was further stratified by disease status into different compartments as shown in Figure 6.

**Figure 6.**
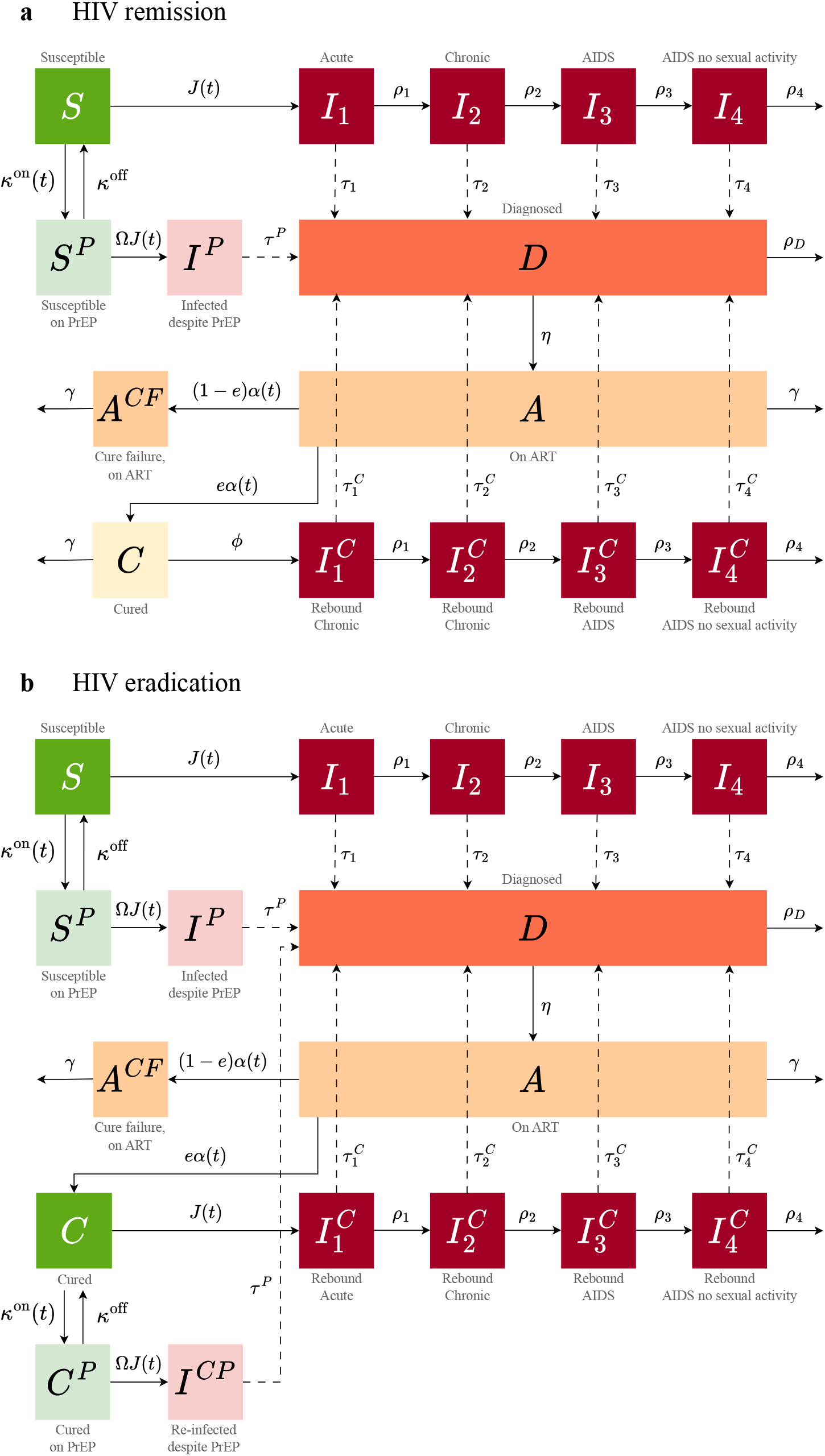
Schematic of the transmission model for two cure scenarios. Transitions are shown for one risk group. Recruitment into and exit from the sexually active population are not shown. A detailed description of the model equations, parameters, and assumptions for (**a**) HIV remission and (**b**) HIV eradication is given in the Supplementary Material.

Susceptible individuals were recruited into the sexually active population at an overall rate of *βN*_0_ and were assigned to risk group *l* with a probability *q*_*l*_, where *N*_0_ is the initial population size, *β* is the rate of entry into the sexually active population, and *q*_*l*_ is the initial fraction of individuals in risk group *l*. Cessation of sexual activity occurred at rate *µ*. In addition, we modeled the immigration of individuals to the Netherlands from abroad who were undiagnosed or already on ART and assigned them to risk group *l* with a probability *Q*_*l*_. Imported undiagnosed infections were assigned to HIV stage *k*, where *k* = 1, 2, 3, 4, with a probability *p*_*k*_.

In each risk group *l*, susceptible individuals acquired HIV at a time-dependent rate *J*_*l*_(*t*) (i.e., the force of infection) through sexual contacts with the infectious individuals. Upon infection, they progressed through four stages: acute (*k* = 1), chronic (*k* = 2), AIDS with severe symptoms (*k* = 3), and AIDS without sexual activity (*k* = 4). These stages were characterized by different progression rates *ρ*_*k*_ and infectivities. Individuals in the last stage died from HIV at rate *ρ*_4_. Infected individuals in stage *k* were diagnosed at rate *τ*_*k*_. Once diagnosed, they either began ART at rate *η* or died from HIV at rate *ρ*_*D*_. The average duration of stay in the infected undiagnosed and diagnosed untreated compartments was assumed to be equal. Individuals on ART left the population at rate *γ* due to a shorter lifespan compared to susceptible individuals. In the Netherlands, the rate of discontinuation of ART among MSM is low, and a variety of next-line ART regimens are available in the event of ART failure. To reflect this, we did not explicitly model ART discontinuation or failure, considering instead that individuals on ART had very low infectivity. Additionally, since individuals in the Netherlands initiate ART promptly upon diagnosis [49] and recently seroconverted MSM reduce their sexual risk behavior following HIV diagnosis [54], we assumed equal infectivity for diagnosed and treated individuals. Susceptible individuals in risk groups *l* = 3, 4 started using PrEP at rates 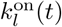 and discontinued it at rate *k*^off^. PrEP reduced the risk of infection with an effectiveness of (1 − Ω). Individuals who acquired HIV while on PrEP had lower infectivity and were diagnosed more rapidly (at rate 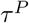) than individuals who did not use PrEP due to 3-monthly HIV testing.

#### Cure scenarios

The schematics of the model for HIV remission and HIV eradication are shown in Figure 6. In both cure scenarios, all individuals on ART, regardless of their risk group, were eligible for a cure. The rollout of the cure was modeled as a time-dependent uptake rate, *α*(*t*), which grew logistically from the start of the rollout in 2026 until it reached a maximum uptake rate, *α*^max^, within 3 years. The efficacy of the cure, *e*, was defined as the proportion of individuals on ART for whom a cure intervention was successful (i.e., who achieved HIV remission or HIV eradication). Individuals who did not achieve a cure or did not take up a cure intervention remained on ART.

Individuals who achieved HIV remission (Figure 6**a**) experienced a viral rebound at a constant rate *ϕ*. We assumed that, upon rebound, individuals entered the chronic HIV stage, similar to what occurs during analytical treatment interruption studies [40, 41], and continued through disease progression at rates *ρ*_*k*_. They were monitored and diagnosed at rates 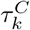,depending on the specific stage *k*, or died from HIV at rate *ρ*_4_. Since the viral reservoir was not completely removed by the intervention, we assumed that individuals in HIV remission died at the same rate as individuals on ART. After achieving HIV eradication (Figure 6**b**), individuals were assumed to become fully susceptible and acquired HIV at the same rate *J*_*l*_(*t*) as naive susceptible individuals. Cured individuals in risk groups *l* = 3, 4 began using PrEP at rates 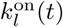 and discontinued it at rate *k*^off^. Upon re-infection, these individuals entered the acute stage, similar to what occurs with superinfection by a different subtype [42], and followed the same disease progression and diagnosis patterns as those in the HIV remission scenario.

#### Model implementation

The model was implemented in R with the RStudio interface (version 2022.12.0+353) using a system of ordinary differential equations for the number of individuals in different compartments. The equations are provided in the Supplementary Material, along with a table describing all model parameters (Table S1) and sensitivity analyses related to key model assumptions.

### Model calibration

The model was calibrated in three steps: (i) sexual partner change rates were estimated from the sexual behavior survey data; (ii) diagnosis rates were estimated from the SHM data on the proportions of diagnoses by time since acquiring HIV; (iii) other parameters were estimated from fitting the model to the SHM data on the annual number of HIV diagnoses and the estimated number of undiagnosed HIV infections from 2017 to 2022.

### Sexual partner change rates

The average number of new sexual partners per year in risk group *l, c*_*l*_, where *l* = 1, 2, 3, 4, was estimated by fitting a probability density function of a Weibull distribution to the sexual behavior data using the maximum likelihood method, as described in detail in [55]. A full explanation of the estimation procedure is reported in the Supplementary Material. The estimated sexual partner change rates are given Table S1 and Figure S2.

In the main analyses presented in Figures 1-6, we assumed that sexual partner change rates remained the same after the introduction of the cure and reflected survey data for the no-cure scenario. In the sensitivity analyses, we relaxed this assumption and considered an increase in sexual risk behavior, as estimated from survey data.

### Diagnosis rates

We fixed the rates of diagnosis in AIDS stages, *τ*_3_ and *τ*_4_, so that the average time until an AIDS diagnosis was 1 month. The rates of diagnosis in the acute and chronic stages, *τ*_1_ and *τ*_2_, were estimated prior to fitting the model to the epidemiological data. To this end, we used a simple stochastic model that simulated the dynamics of disease progression and diagnosis, aiming to reproduce the proportions of diagnoses within 6 months, between 6 to 12 months, and more than 12 months since acquiring HIV, based on the SHM data [2]. The detailed estimation procedure is described in the Supplementary Material. The estimated diagnosis rates are given in Table S1 and Figure S4.

### Other estimated parameters

The estimated parameters were the probability of transmission per sexual partner, *λ*; the parameter describing mixing between risk groups, *ω*; the relative infectivity of diagnosed and treated individuals, *ε*; the number of undiagnosed individuals immigrating to the Netherlands from abroad per year *M*_*I*_; the initial number of undiagnosed individuals for model burn-in, *U*_0_; and the probability of initial and imported HIV infections being assigned to risk group *l, Q*_*l*_, where *l* = 1, …, 4. These parameters were estimated by fitting the model to the SHM data on new HIV diagnoses and the estimated number of undiagnosed HIV infections from 2017 to 2022, using an Approximate Bayesian Computation approach based on Latin Hypercube Sampling. The detailed procedure for obtaining the posterior distributions of all estimated parameters is presented in the Supplementary Material. The estimated parameters are given in Table S1 and Figure S5. The remaining model parameters were fixed based on the literature (Table S1).

### Varied cure parameters

The parameters varied in both cure scenarios were efficacy, *e*, and maximum annual uptake percentage, 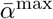.In the HIV remission scenario, we also varied the average time until viral rebound, 1*/ϕ*. The maximum annual uptake percentage was calculated as 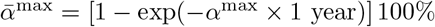,where *α*^max^ is the maximum uptake rate per year used in the model equations.

Monitoring strategies were defined by varying the diagnostic delay for rebounds/re-infections in infection stage 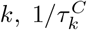:(i) No monitoring, a diagnostic delay as estimated under the current standard of care: 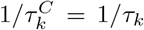 for all *k* with *τ*_*k*_ values taken from the estimated posterior distribution, resulting in an average (across all HIV stages) diagnostic delay of 28 months. (ii) PrEP-like monitoring: 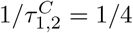 (acute and chronic stages) and 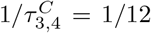 (AIDS stages), resulting in an average diagnostic delay of 3 months, recommended for PrEP users in the Netherlands. (iii) Frequent monitoring: 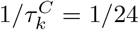 for all *k*, yielding an average diagnostic delay of 0.5 months.

### Model outcomes

The model outcomes were annual new HIV infections (primary infections in naive individuals) and the change in cumulative HIV infections compared to the no-cure scenario from 2026 to 2034. For HIV remission, we estimated annual and cumulative viral rebounds; for HIV eradication, we estimated annual and cumulative re-infections (secondary infections in cured individuals) over the same period. For HIV remission, HIV prevalence was calculated as the ratio of the sum of individuals who are infected, diagnosed, treated, and in remission and the total population size. Individuals in remission were included in the prevalence calculation because their HIV reservoir is not completely removed. For HIV eradication, the compartments considered for HIV prevalence calculation were infected, diagnosed, and treated. Cure coverage was calculated as the proportion of cured individuals among all eligible (all treated and cured compartments).

### Sensitivity analyses

In the sensitivity analyses provided in the Supplementary Material, we explored the impact on model projections of (i) the year of cure introduction, (ii) risk compensation following cure introduction, (iii) infectivity of individuals after rebound and re-infection, (iv) the underlying trend in HIV incidence, (iv) infectivity of diagnosed individuals, (v) duration of survival of individuals achieving HIV remission, and (vi) susceptibility to re-infection of individuals achieving HIV eradication.

## Data Availability

All data produced in the present study will be available upon publication.

## Data availability

The data used in this study will be avaible upon publication at https://github.com/alfredodebellis/HIVcure.

## Code availability

The codes reproducing the results of this study will be avaible upon publication at https://github.com/alfredodebellis/HIVcure.

## Additional information

The Supplementary Material provides additional figures, tables, and further details of this study.

## Acknowledgements

The authors gratefully acknowledge funding from Aidsfonds Netherlands (grant number P53902) and the Netherlands Organization for Scientific Research (NWO), SPIRAL project KICH2.V4P.AF23.001. We extend our gratitude to the survey participants and thank the staff of the Amsterdam Cohort Studies, the AGEhIV Cohort Study, and the infectious diseases outpatient clinic at the University Medical Center Utrecht for their assistance in participant recruitment. Special thanks to Thijs Albers, Bertus Tempert, and Renee Finkenflügel from the Dutch HIV Association. We also appreciate the valuable discussions with Franco Romero Gonzalez, Maartje Dijkstra, Maartje Basten, Laura de Groot, Sigrid Vervoort, Berend van Welzen, Udi Davidovich, Amy Matser, Myrthe Verburgh, Pythia Nieuwkerk, Godelieve de Bree, and Janneke Heijne.

## Author contributions

G.R. conceived and supervised the study. A.D.B. and G.R. developed the mathematical model. A.D.B., G.R., and G.G. developed the fitting procedure. K.R. and A.v.S. collected and provided data. K.R., P.R., M.S.v.d.L., A.D.B., M.N., and G.R. developed cure scenarios. M.W. performed a literature review. A.D.B. conducted analyses and prepared figures. A.D.B., M.W., and G.R. wrote the manuscript. All authors contributed to the interpretation of the results, edited the manuscript, and gave final approval for publication.

## Correspondence

Correspondence and material requests should be addressed to Dr. Ganna Rozhnova (Julius Center for Health Sciences and Primary Care, University Medical Center Utrecht, P.O. Box 85500 Utrecht, The Netherlands) and Alfredo De Bellis (Health Emergencies Center, Fondazione Bruno Kessler, P.O. Box 38123 Trento, Italy).

## Supplementary Material

### 1 Model equations with cure

#### 1.1 HIV remission

The equations for the numbers of men who have sex with men (MSM) in risk group *l, l* = 1, …, 4, who are susceptible (*S*_*l*_), susceptible on pre-exposure prophylaxis (PrEP) (*S*^*P*^), infected but not diagnosed (*I*_*lk*_) in stage *k, k* = 1 (acute), 2 (chronic), 3 (AIDS with onset of severe symptoms), 4 (AIDS without sexual activity), infected while on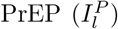,diagnosed (*D*_*l*_), on antiretroviral treatment (ART) (*A*_*l*_), in remission — virally suppressed below transmissible levels without ongoing ART (*C*_*l*_), not effectively suppressed after HIV remission strategy and thus still on ART 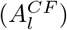,infectious due to viral rebound 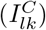 read as follows

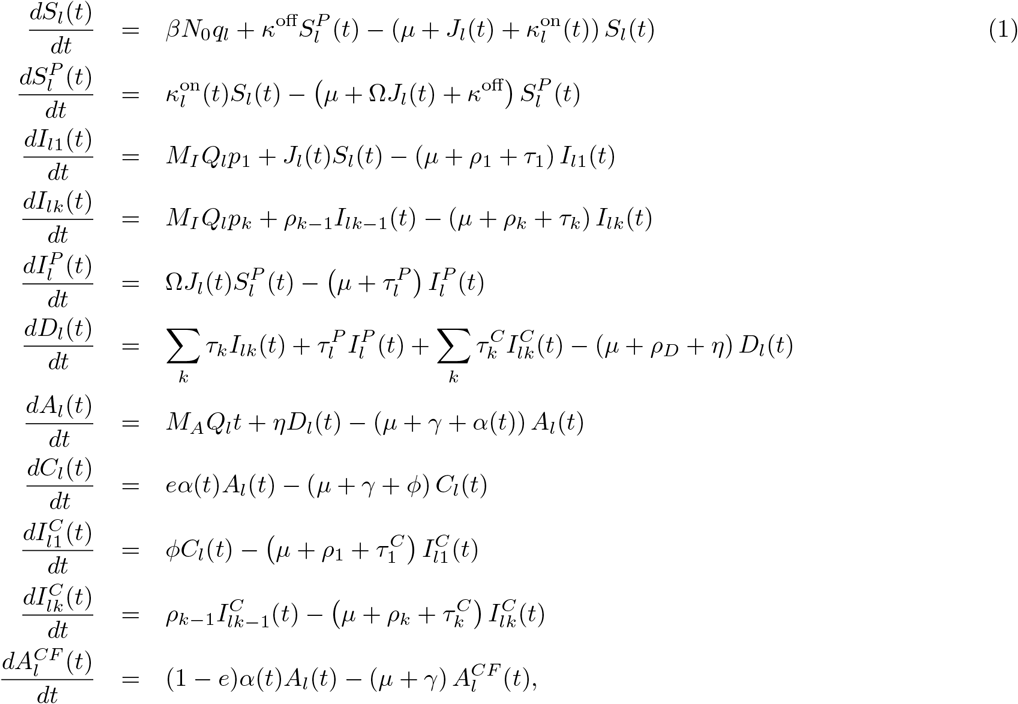

where *k* = 2, 3, 4 and *l* = 1, …, 4.

The force of infection in risk group *l* is written as

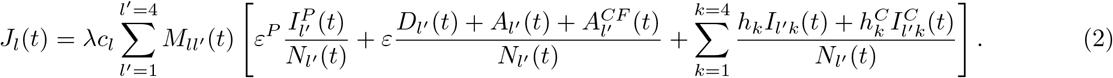

In Equation 2, *N*_*l*_(*t*) is the population size in risk group *l*

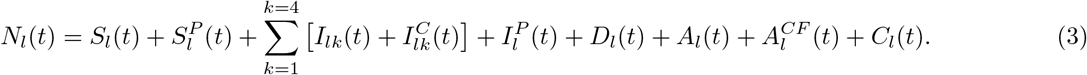

Following [1–4], we define the 4 *×* 4 mixing matrix 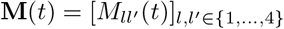,where the elements 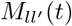 represent mixing between susceptible individuals in risk group *l* and infectious individuals in risk group *l*^*′*^, as follows

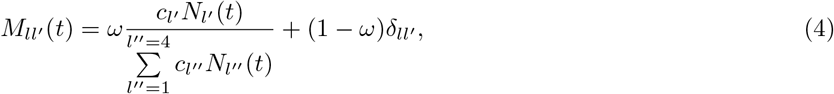

where 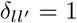 and 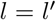 otherwise. The mixing parameter 0 ≤ *ω* ≤ 1 describes the degree of assortative mixing by risk group. When *ω* = 0, mixing between risk groups is fully assortative (like with like), whereas *ω* = 1 indicates fully proportionate mixing. Equation 4 implies that a proportion (1 − *ω*) of partnerships are formed exclusively within the same risk group *l* = *l*^*′*^, while the remaining proportion *ω* of partnerships is formed across all risk groups (*l*^*′*^ = 1, …, 4) proportionally to the number of partnerships offered by each group.

The description of the model parameters is given in Table S1.

#### 1.2 HIV eradication

The equations for the numbers of MSM in risk group *l, l* = 1, …, 4, who are naive susceptible (*S*_*l*_), cured — susceptible after complete removal of HIV (*C*_*l*_), naive susceptible on PrEP 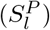,cured on 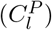,primarily infected (*I*_*lk*_), re-infected after a cure (*I*^*C*^) in stage *k, k* = 1 (acute), 2 (chronic), 3 (AIDS with onset of severe symptoms), 4 (AIDS without sexual activity), primarily infected while on PrEP (*I*^*P*^), re-infected after a cure while on PrEP 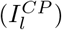,diagnosed (*D*_*l*_), on ART (*A*_*l*_), not effectively cured after HIV eradication strategy and thus still on ART 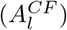 are given by

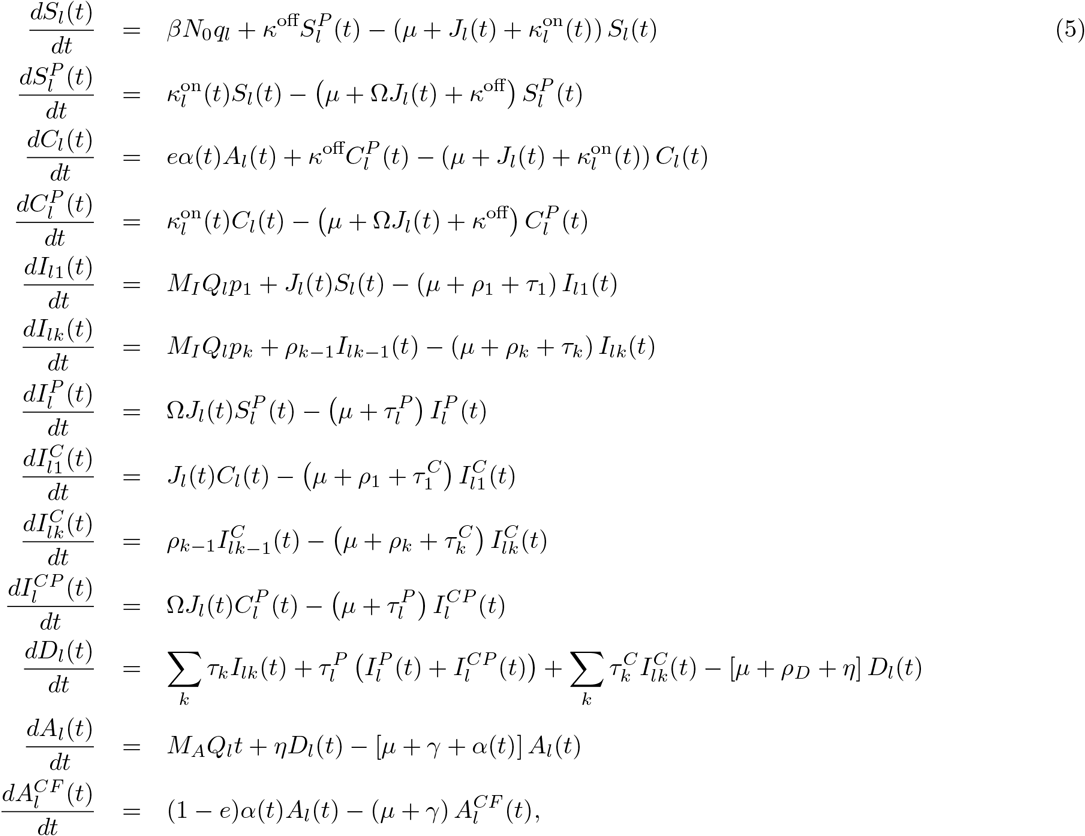

where *k* = 2, 3, 4 and *l* = 1, …, 4.

The force of infection in risk group *l* is written as

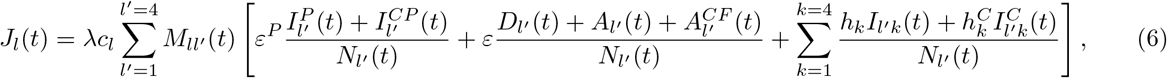

where *N*_*l*_(*t*) is the population size in risk group *l*

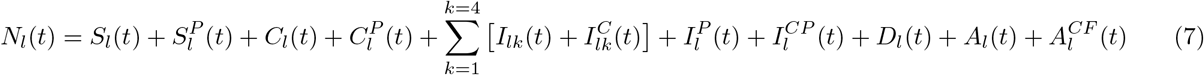

and the mixing matrix 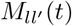 is given by Equation 4.

The description of the model parameters is given in Table S1.

### 2 Model calibration and validation

#### 2.1 Estimation of sexual partner change rates

The model explicitly accounted for risk heterogeneity in sexual activity by stratifying the MSM population into four risk groups based on the average number of new sexual partners per year (also termed ‘partner change rate’), *c*_*l*_, where the subscript *l* = 1, …, 4 denotes risk groups [4]. Here, *l* = 1 represents the group with the lowest number of partners and *l* = 4 represents the group with the highest number of partners. Individuals remained in the same risk group. Although the model can accommodate any number of risk groups, we focused on four risk groups, following a previous study on the impact of PrEP on HIV dynamics among MSM in the Netherlands [4]. From this study, we adopted the initial population fractions in each risk group, *q*_*l*_, where 0 *< q*_*l*_ ≤ 1 for *l* = 1, …, 4 and 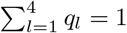 (Table S1). Specifically, the population fractions in the groups with the lowest and the highest numbers of new sexual partners per year were *q*_1_ = 0.45 and *q*_4_ = 0.07, respectively.

The average number of new sexual partners per year in risk group *l, c*_*l*_, where *l* = 1, …, 4, was estimated using sexual behavior data from a cross-sectional online survey that aimed to assess the expected impact of an HIV cure on quality of life, sexual satisfaction, stigma, and sexual and preventive behaviors among people with HIV and key populations in the Netherlands. Details on the survey can be found in [5], and the questionnaire is available at https://github.com/alfredodebellis/HIVcure. The study received ethical approval from the University Medical Center Utrecht (UMC Utrecht) ethics committee (20-546/C).

Participants were eligible if they were Dutch or English-speaking, 18 years or older, living with HIV, or part of key populations such as MSM, and provided informed consent. Recruitment occurred from October 2021 to June 2022, drawing from the Amsterdam Cohort Studies (ACS) [6], the AGEhIV Cohort Study [7], the infectious diseases outpatient clinic of the UMC Utrecht, the Dutch HIV Association [8], Stichting ShivA [9], Grindr [10], and the Public Health Services of Amsterdam, Rotterdam, Zuid-Limburg, Groningen, Utrecht, Gelderland-East, and Brabant-South East. Data were collected using REDCap electronic data capture tools hosted by the UMC Utrecht and the ACS. Questions were tailored to ensure participants only saw those relevant to them.

From this survey, we extracted and analyzed responses from *n* = 529 MSM participants who provided information on their number of casual and steady partners in the last six months and condom use. Questions were asked three times, assuming: (i) the current situation (i.e., no cure available); (ii) an HIV remission scenario becomes available; and (iii) an HIV eradication scenario becomes available. For (i) the current situation, participants were asked: “How many different casual sex partner(s) did you have anal and/or vaginal intercourse with in the last 6 months?” and “How often did you use condoms during anal and/or vaginal intercourse with your casual sex partner(s) in the last 6 months?”. Scenarios (ii) and (iii) gauged anticipated changes in sexual behavior if a cure became available. Participants responded to: “How many different casual sex partner(s) would you have anal and/or vaginal intercourse with?” and “How often would you use condom(s) during anal and/or vaginal intercourse with your casual partner(s)?” Similar questions were asked about steady partners.

Condom use responses were coded as a binary variable (0 = always, 1 = not always), categorizing all MSM who reported condomless anal intercourse in the last six months into the ‘not always’ group. This binary variable was applied as a multiplier to the number of partners, so MSM who consistently used condoms were effectively assigned a partner count of zero. To obtain the total number of new sexual partners, we adjusted the number of casual partners by adding zero, one, or two steady partners per individual based on multinomial probabilities 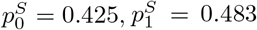, and 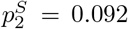, estimated from data for the current situation among MSM recruited through the ACS [6].

Empirical cumulative histograms from the survey data are shown as open circles in Figure S2 for (i) the current situation (or no-cure scenario), (ii) an HIV remission scenario, and (ii) an HIV eradication scenario. We estimated partner change rates in risk group *l, c*_*l*_, *l* = 1, …, 4, by fitting a Weibull probability density function to the histograms using the maximum likelihood method [3]. The fitted Weibull distributions are shown as blue lines in Figure S2, with their estimated shape and scale parameters listed in the legend of each panel. To calculate the average number of new sexual partners per risk group, *c*_*l*_, we used intervals defined by the initial fractions of the population in risk groups, *q*_*l*_, *l* = 1, …, 4. In this calculation, the proportions of the population, *q*_*l*_, in each risk group were kept the same for all three scenarios. The vertical dashed lines in Figure S2 indicate the intervals defining partner change rates for each risk group. The results obtained represent the rates per six months. We multiplied the averages by 1.5 to obtain the number of new partners per year, because some partnerships are likely to last more than six months [11, 12]. The average numbers of new partners in risk group *l* for all three scenarios are reported in Table S1.

Because all inferred changes in sexual behavior post-cure are hypothetical, partner change rates in the main analyses were fixed at values estimated for the current situation (Figure S2**a**). In sensitivity analyses, this assumption was relaxed, and we used values estimated in scenarios shown in Figure S2**b** and S2**c**. Further details are provided in Section 3.3. We also verified that partner change rates inferred in other studies (e.g., [4]) result in similar model fits to epidemiological data.

#### 2.2 Estimation of diagnosis rates

We implemented a simple stochastic model that simulates the state progression of *Z* = 1000 individuals initialized at the acute stage. We sampled *N*_*τ*_ = 10000 values of diagnosis rates for acute stage, *τ*_1_, and chronic stage, *τ*_2_, uniformly distributed in the intervals [1/12, 2] 1/year and [1/12, 1] 1/year, respectively. We set diagnosis rates for the AIDS stages 3 and 4, *τ*_3_ and *τ*_4_ respectively, such that the average waiting time before being diagnosed with AIDS is 1 month (Table S1). Then, for each sampled pair, we ran the model described in Pseudocode 1. We define a relative error threshold *ϵ* = 10% and accept all the pairs such that the proportions of diagnoses within 6 months, between 6 to 12 months, and more than 12 months since HIV infection match (with a tolerance given by *ϵ*) data from the Dutch HIV Monitoring Foundation (Stichting hiv monitoring, SHM) [13]. The reconstructed proportions of diagnoses are reported in Figure S3. The distributions of *τ*_1_ and *τ*_2_ are shown in Figure S4.

##### Algorithm 1 Pseudocode for the stochastic model to build the distribution of diagnostic delays

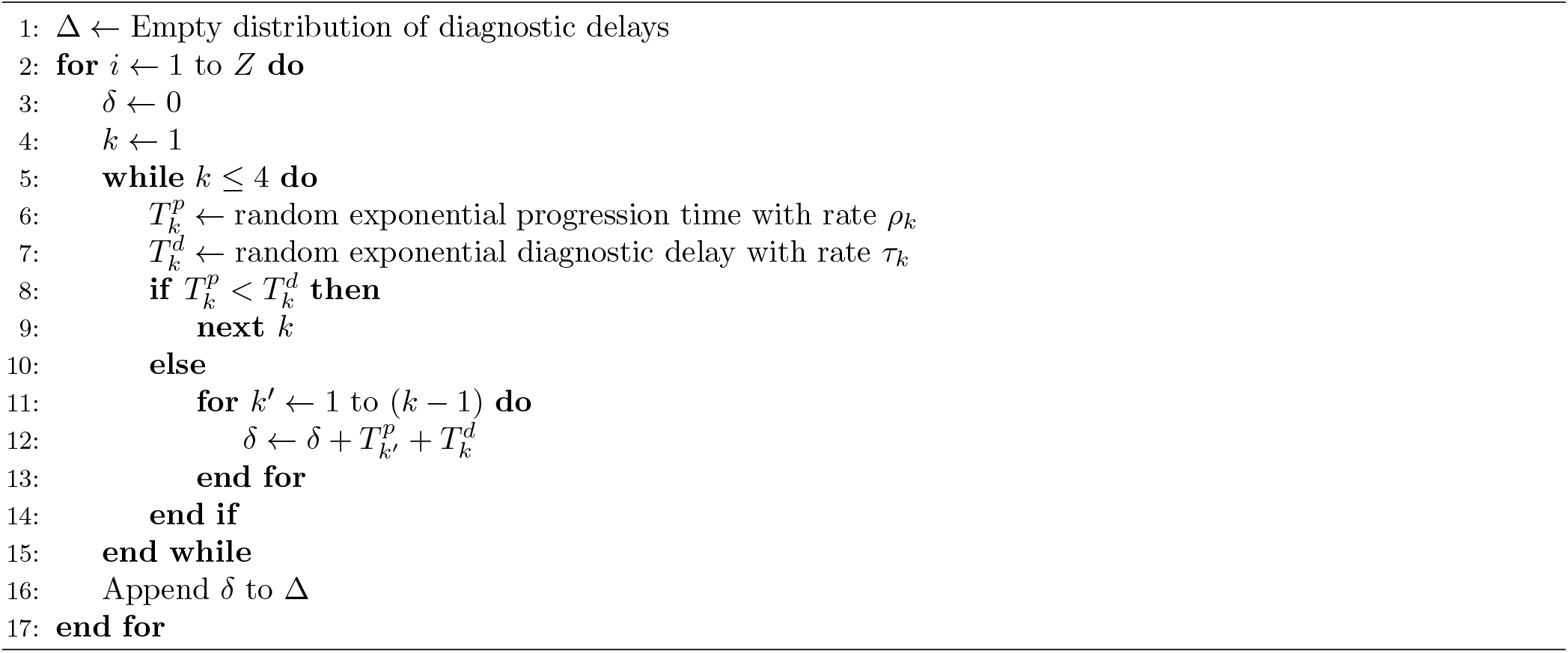

#### 2.3 Estimation of other parameters

The estimated parameters were the probability of transmission per sexual partner, *λ*; the parameter describing mixing between risk groups, *ω*; the relative infectivity of diagnosed and treated individuals, *ε*; the number of undiagnosed individuals immigrating to the Netherlands from abroad per year *M*_*I*_; the initial number of undiagnosed individuals, *U*_0_; and the probability of initial and imported HIV infections being assigned to risk group *l, Q*_*l*_, where *l* = 1, …, 4.

We used an Approximate Bayesian Computation (ABC) framework based on Latin Hypercube Sampling (LHS) to estimate these parameters. The procedure consists of two sequential steps. First, we sampled from *L* = 40000 parameters sets, generated through the LHS based on the prior ranges reported in Table S2, for which the condition *Q*_1_ *< Q*_2_ *< Q*_3_ *< Q*_4_ holds. After normalizing all the *Q*_*l*_ such that ∑ _*l*_ *Q*_*l*_ = 1, we sampled parameters from the generated list and simulated the epidemic dynamics for each set. The performance of each simulation was evaluated by comparing the simulated new HIV diagnoses per year and the number of undiagnosed individuals with the SHM data time series [13]. We accepted a parameter set if it met the following criteria: i) the simulated yearly diagnoses fell within 10% of the observed data points, and ii) the annual number of undiagnosed HIV infections remained within the uncertainty intervals of the observed data throughout the calibration period (2017-2022). Additionally, we constrained the estimated annual number of undiagnosed HIV infections due to immigration of individuals to the Netherlands from abroad to be below the upper bound of the 2022 HIV incidence (150 new HIV infections as estimated by SHM [13]).

The model fit is reported in Figure 1. The empirical distribution of accepted parameters and the Pearson correlations between parameters are shown in Figure S5 and Figure S6, respectively.

#### 2.4 PreP use and immigration with HIV from abroad

Based on observed data, we assume a linear increase with time in the annual number of individuals on PrEP and the annual number of individuals on ART immigrating to the Netherlands from abroad (Figure S1), while undiagnosed infected individuals immigrate at a constant rate *M*_*I*_. We then fitted two linear models to estimate the lines’ slopes, *M*_*A*_ and *M*_*P*_, to inform the model variables, *A*(*t*) and *S*^*P*^ (*t*). While *M*_*A*_ enters directly the equations of the model for the compartment *A*(*t*) as *M*_*A*_*t* (see Equations 1 and 5), further assumptions should be made for PrEP users. Data on the number of PrEP users in the national PrEP program from the national sexually transmitted infections surveillance database [14], available from June 2019 to April 2022, were taken from [15]. However, since pilot PrEP studies began in 2015, we modeled PrEP uptake starting from that year. Hence, we divided the period into two, namely 2015-2019 and 2019-2021, and estimated two different slopes for these periods, 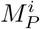 and 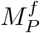, respectively. The national program provides PrEP to MSM at high risk of HIV acquisition. We modeled this by making only risk groups 3 and 4 eligible for PrEP and weighted 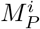 and 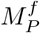 by the population fractions *q*_3_ and *q*_4_, resulting in four different values: 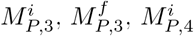 and 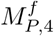. To apply the linearity assumption for PrEP users in our model, we use the equation for the derivative of 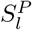 and focus on the case where the outflow from the compartment, aside from PrEP dropout, is negligible:

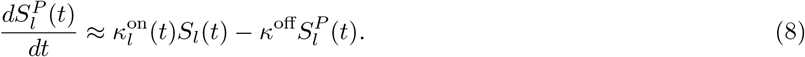

A linear growth of *S*^*P*^ translates into a constant derivative, *dS*^*P*^ */dt* ≈ *M*_*P*_. Substituting this into Equation 8, we find that the PrEP uptake rate for risk groups *l* = 3, 4 has the following form:

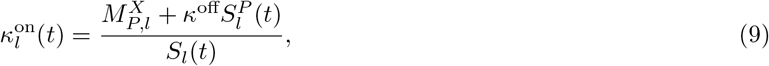

where *X* = *i* before 2019 and *X* = *f* after. We assume that, after 2022, the number of PrEP users remains at a maximum capacity of *N* ^max^ = 10^4^ because the capacity of the state-funded PrEP program in the Netherlands is limited.

#### 2.5 Model validation

The model was validated on the number of PrEP users and the annual number of individuals on ART immigrating to the Netherlands from abroad (Figure S1**a** and Figure S1**b**). We also validated our model calibration using ART coverage among all MSM with HIV over time (Figure S1**c**). The model correctly captured the trend in ART coverage but slightly overestimated the data, potentially because it did not account for individuals lost to follow-up due to moving abroad or other reasons. An additional validation of our model is reported in Figure S7, where we show that the average proportions of infected individuals per risk group remain almost constant throughout the time window of the simulation.

### 3 Sensitivity analyses

We explored the impact on model projections of (i) the year of cure introduction, (ii) risk compensation following cure introduction, (iii) infectivity of individuals after rebound and re-infection, and (iv) the underlying trend in HIV incidence. Additional sensitivity analyses are reported at the end of this section.

#### 3.1 Time of cure introduction

The year of cure introduction is highly uncertain. In the main analyses, the cure was assumed to be introduced in 2026. Figure S8 and Figure S9 show projections of HIV dynamics if the cure is introduced in 2030. These figures can be compared directly with Figure 2 (for HIV remission) and Figure 4 (for HIV eradication).

#### 3.2 Infectivity of rebounds/re-infections

Based on data from ART interruption studies, we assumed in our main analyses of the HIV remission scenario that the infectivity of individuals after viral rebound would be equal to that observed during the chronic stage of HIV infection [16, 17]. Since chronic-stage infectivity is lower than acute-stage infectivity (Table S1), the projections for HIV remission shown in Figure 2 represent the most optimistic scenario. In Figure S10, we present alternative projections of HIV dynamics using the same parameters as in Figure 2, but assuming that the infectivity of individuals after viral rebound matches that of the acute stage. Unlike the results in Figure 2**a**, these projections show an increase in new HIV infections compared to the no-cure scenario, even when the time to rebound is as long as six years (Figure S10**a**). The high infectivity could undermine the effectiveness of the transient HIV remission strategy and should be carefully considered when developing a cure.

Conversely, in the HIV eradication scenario, we assumed that the infectivity of individuals re-infected after HIV eradication would resemble the acute stage of HIV infection, as observed for superinfection by a different HIV subtype [18] or re-infection with hepatitis C virus [19, 20]. In Figure S11, we present projections of HIV dynamics under the assumption that the infectivity of the initial stage following re-infection equals that of the chronic stage. For all cure uptakes considered, these estimates were similar to those shown in Figure S4. Thus, the overall conclusions regarding the HIV eradication strategy remained consistent across different assumptions about the infectivity of re-infections.

#### 3.3 Behavioral changes after cure

In the main analyses shown in Figures 2-5, partner change rates were fixed at values estimated from the survey data for (i) the current situation without a cure. In the sensitivity analyses, we relaxed this assumption and instead used partner change rates estimated from the survey data assuming that (ii) an HIV remission scenario becomes available and (iii) an HIV eradication scenario becomes available. The estimated ‘survey’ partner change rates for these three scenarios were (Table S1): (i) *c*_1_ = 0.52, *c*_2_ = 3.81, *c*_3_ = 11.16, *c*_4_ = 28.56 1/6 months; (ii) *c*_1_ = 1.03, *c*_2_ = 6.42, *c*_3_ = 17.15, *c*_4_ = 40.02 1/6 months; (iii) *c*_1_ = 0.90, *c*_2_ = 5.90, *c*_3_ = 16.25, *c*_4_ = 39.04 1/6 months. The proportions of the population in each risk group, *q*_*l*_, were kept constant across all three scenarios: *q*_1_ = 0.45, *q*_2_ = 0.35, *q*_3_ = 0.13, *q*_4_ = 0.07 (Table S1). To simulate behavioral changes after a cure, we assumed that partner change rates gradually transition over a period of 3 years from pre-cure levels (scenario (i)) to post-cure levels (either (ii) for the HIV remission scenario or (iii) for the HIV eradication scenario). This transition period aligned with the increase in the rate of cure uptake. We also examined the impact of reduced behavioral changes, where post-cure partner change rates were set as the average of pre-cure (i) and post-cure rates (ii) or (iii) estimated from the survey: (i) *c*_1_ = 0.52, *c*_2_ = 3.81, *c*_3_ = 11.16, *c*_4_ = 28.56 1/6 months; (ii) *c*_1_ = 1.03, *c*_2_ = 6.42, *c*_3_ = 17.15, *c*_4_ = 40.02 1/6 months; (iii) *c*_1_ = 0.90, *c*_2_ = 5.90, *c*_3_ = 16.25, *c*_4_ = 39.04 1/6 months.

Figure S12 illustrates HIV dynamics with behavioral changes after the introduction of the transient HIV remission scenario (Figure S12**a** and **b**) and the HIV eradication scenario (Figure S12**c** and **d**). In these analyses, the time to viral rebound was set at 6 years, with a 3-month diagnostic delay for individuals who experienced rebound or re-infection. Cure uptake and efficacy were fixed at 90%.

In the transient HIV remission scenario (Figure S12**a**), the number of new HIV infections in 2034 could exceed that of the no-cure scenario if behavioral changes were introduced. Since behavioral changes temporarily increased HIV infections, we also calculated cumulative infections over the 8 years of the simulation corresponding to Figure S12**a**. Specifically, cumulative HIV infections over 8 years increased by 9% relative to the no-cure scenario (orange lines) when survey partner change rates were used (purple line). Without behavioral changes (blue line) or with reduced behavioral changes where post-cure partner change rates were set as the average of pre-cure and post-cure rates (black line), cumulative HIV infections over 8 years decreased by 27% and 9%, respectively, relative to the no-cure scenario. Despite varied levels of behavioral changes, the number of new rebounds per 100,000 individuals per year remained the same (Figure S12**b**). Therefore, behavioral changes leading to risk compensation could further limit the effectiveness of the transient HIV remission scenario and potentially increase cumulative HIV infections even further relative to the no-cure scenario.

In the HIV eradication scenario (Figure S12**c**), the number of new HIV infections in 2034 remained lower than in the no-cure scenario, even with the introduction of behavioral changes. Specifically, cumulative HIV infections over 8 years decreased by 41% relative to the no-cure scenario (orange lines) when using survey partner change rates (purple line). Without behavioral changes (green line) or with reduced behavioral changes (black line), cumulative HIV infections over 8 years decreased by 58% and 49%, respectively, relative to the no-cure scenario.

#### 3.4 Underlying trend in HIV incidence

Our projections presented in the main text were not intended as precise forecasts, as future changes in the national PrEP program, sexual behavior or migration trends could influence the quantitative predictions for the no-cure scenario, potentially leading to an increase in HIV incidence. Figures S13**a,b** and S13**c,d** illustrate HIV dynamics under the HIV remission and HIV eradication scenarios, respectively, assuming an increase in HIV incidence in the no-cure scenario starting in 2022. In these analyses, the time to viral rebound was set at 6 years, with a 3-month diagnostic delay for individuals who experienced rebound or re-infection. Cure uptake and efficacy were fixed at 90%. The post-2022 increase in HIV incidence was implemented as *λ*_incr_(*t*) = *λ* + *ψ*(*t* − *T*_*L*_), where *ψ* = 0.001, *T*_*L*_ = 2022, and *λ* is based on 100 samples from the posterior distribution. These sensitivity analyses show that the qualitative insights into the impact of cure scenarios relative to the no-cure scenario hold despite changes in the underlying trend in HIV incidence.

#### 3.5 Infectivity of diagnosed individuals

Since individuals in the Netherlands initiate ART promptly upon diagnosis [21] and recently seroconverted MSM reduce their sexual risk behavior following HIV diagnosis [22], we assumed in the main analyses that diagnosed but untreated individuals and those on ART have equal infectivity. Although this assumption may be somewhat unrealistic, its impact is minor, as the average duration in the diagnosed compartment prior to viral suppression is short (i.e., 1*/η* = 1.5 months [21]). To test the robustness of this assumption, we assigned a higher infectivity for diagnosed but untreated individuals, equivalent to the mean infectivity of undiagnosed individuals, and found that our results remained consistent.

#### 3.6 Duration of survival of individuals achieving HIV remission

In the main analyses of the HIV remission scenario, we assumed that individuals achieving HIV remission would have a survival duration equivalent to those on stable ART (1*/γ* = 61 years). Hypothetically, there may be no additional HIV-related mortality for individuals in remission. Setting *γ* = 0 for these individuals had minimal impact on our results.

#### 3.7 Susceptibility of individuals achieving HIV eradication

In the main analyses of the HIV eradication scenario, we assumed that individuals who achieved HIV eradication would be fully susceptible to re-infection. A scenario in which individuals are fully protected from re-infection is equivalent to the sustained HIV remission scenario analyzed in the main text. For cases of partial susceptibility, the results fall between those of the HIV eradication and sustained HIV remission scenarios.

## 4 Supplementary Tables

**Table S1.**
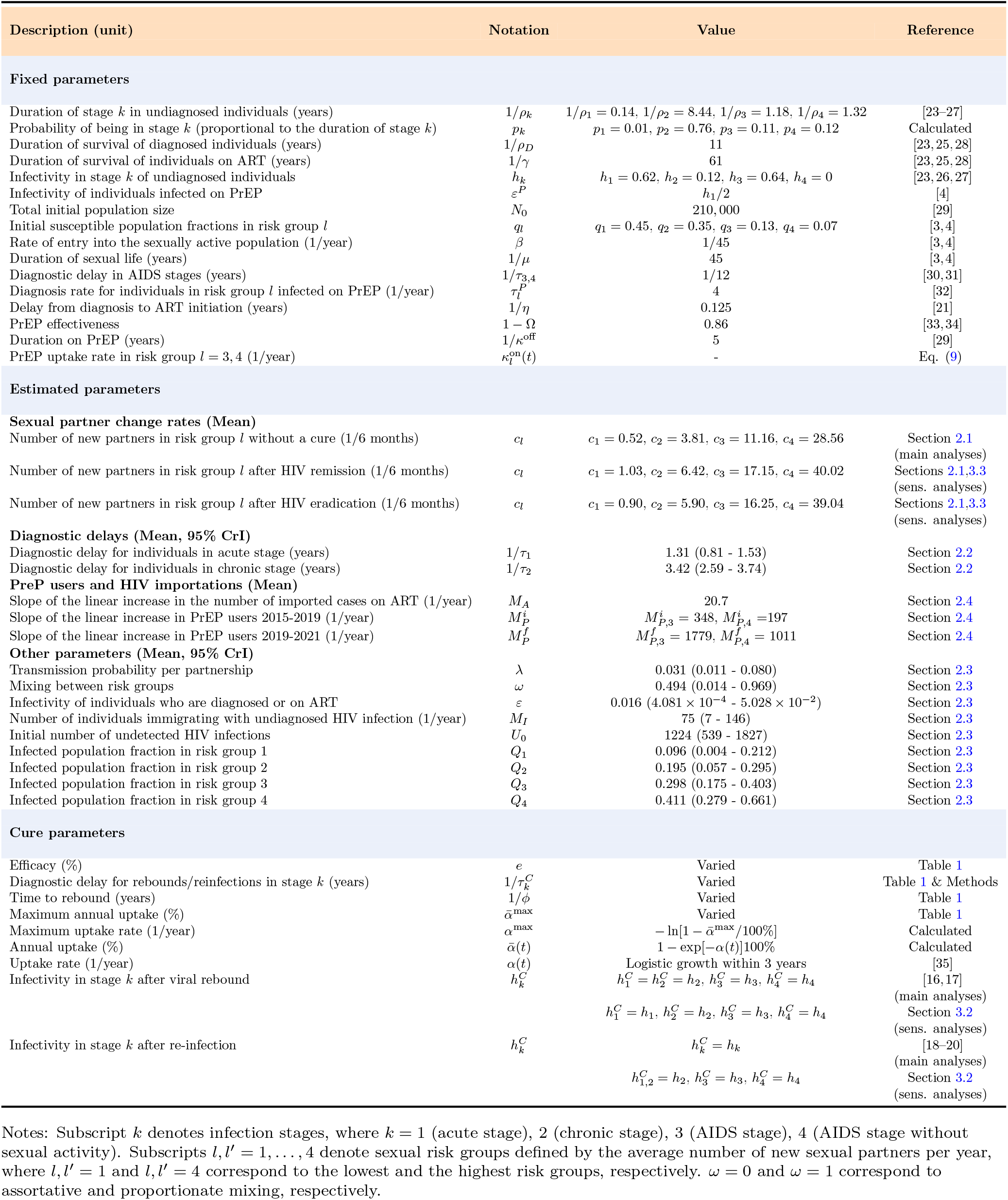
Summary of the model parameters.

**Table S2.** Uniform priors for the parameters estimated in the model.

| Parameter | Interval |
| --- | --- |
| $\tau_1$ | [0.01, 2] |
| $\tau_2$ | [0.01, 1] |
| $\lambda$ | [0.01, 0.15] |
| $\omega$ | [0, 1] |
| $\varepsilon$ | [0, 0.06] |
| $M_I/(\beta N_0)$ | [0, 0.05] |
| $U_0$ | [0, 2000] |
| $Q_l, l = 1, \dots, 4$ | [0, 1] |

## 5 Supplementary Figures

**Figure S1.**
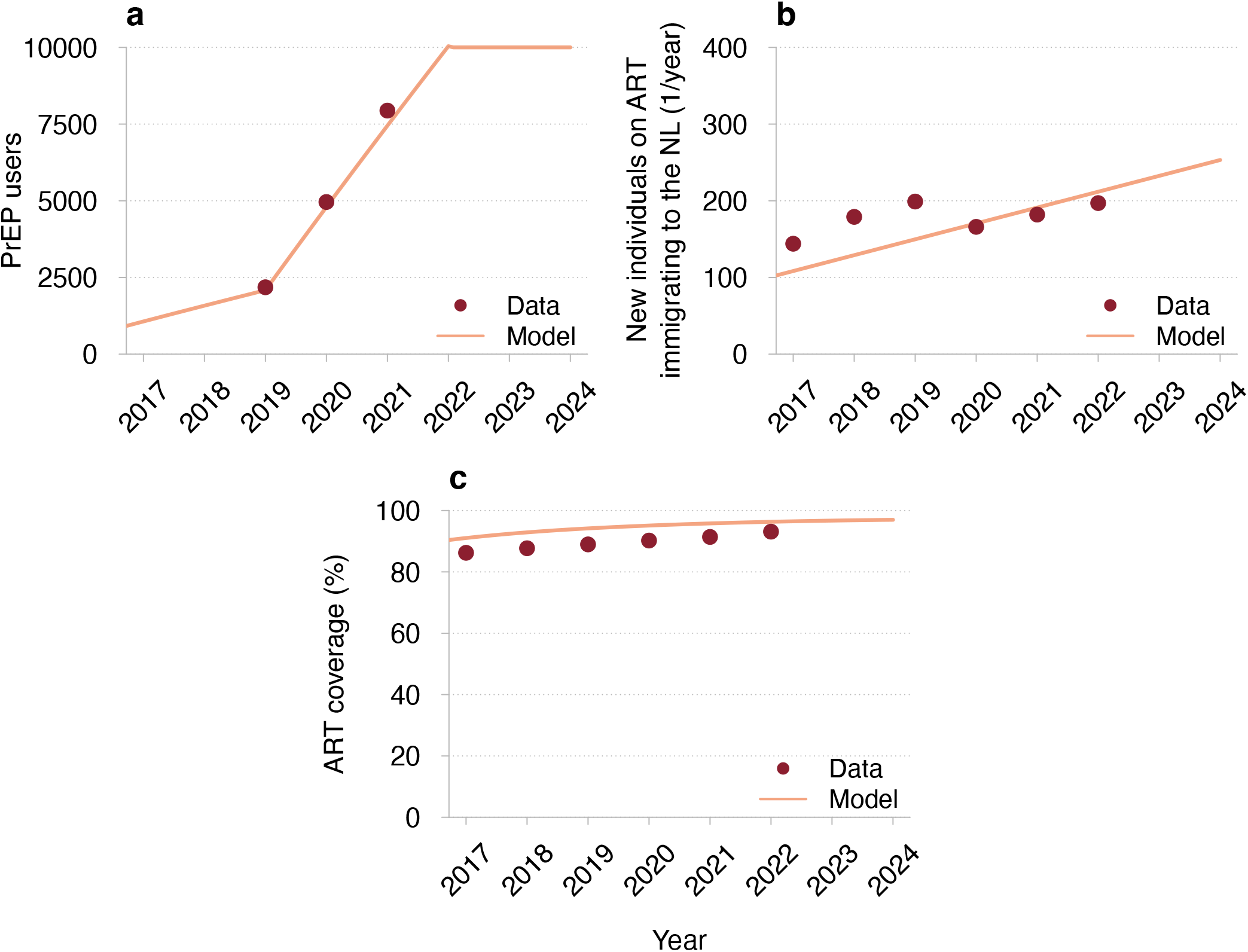
Model validation against HIV surveillance data for MSM in the Netherlands. (**a**) Number of individuals on PrEP, (**b**) number of individuals on ART immigrating to the Netherlands from abroad, and (**c**) ART coverage among all MSM with HIV. The mean trajectories estimated from the model are shown as orange lines. The orange-shaded regions correspond to 95% credible intervals based on 100 samples from the joint posterior parameter distribution. The data (red dots) are data on the number of PrEP users in the national PrEP program from the national sexually transmitted infections surveillance database taken from [15] for (**a**) and from the SHM report [13] for (**b**) and (**c**).

**Figure S2.**
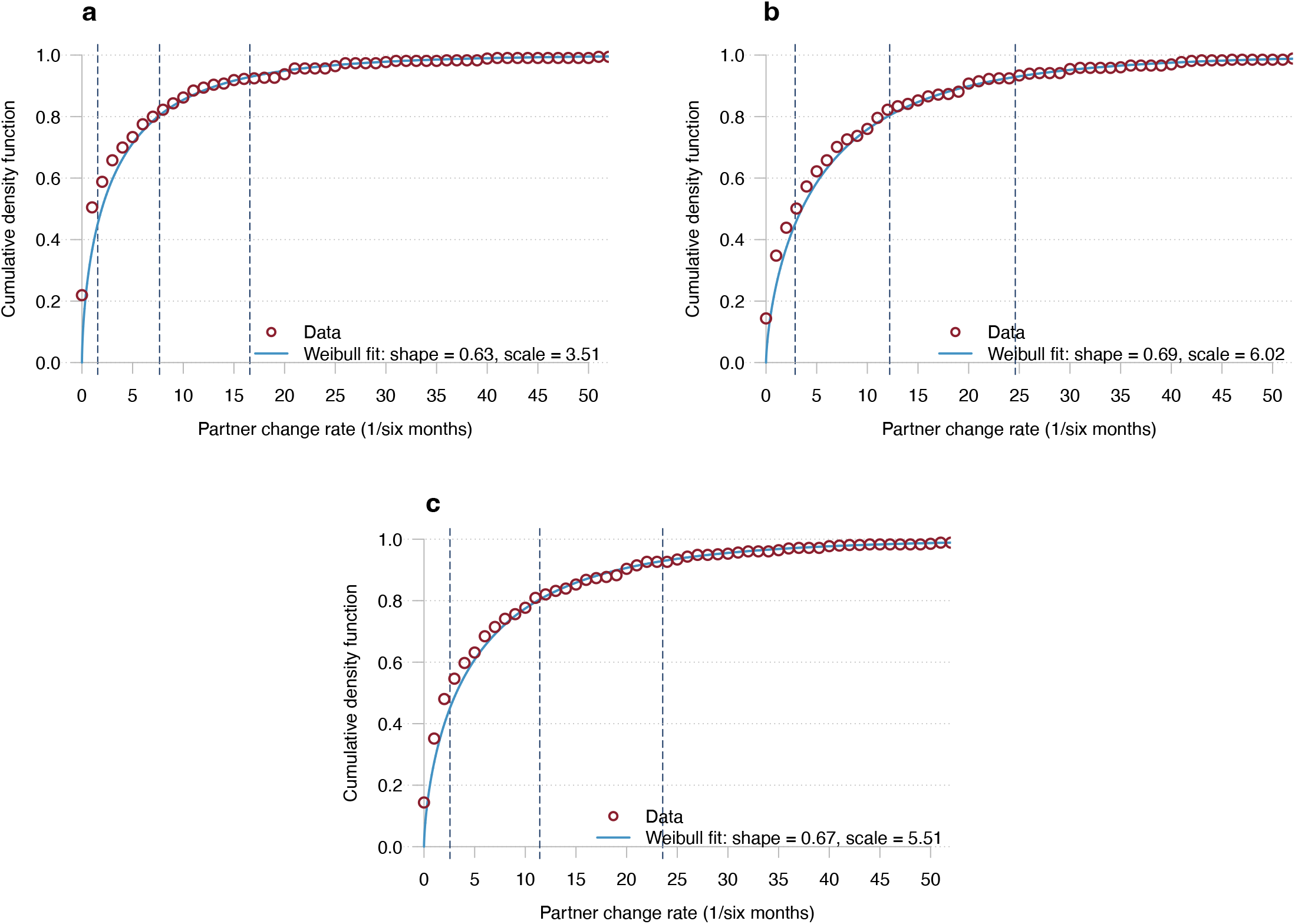
Cumulative probability density function of sexual partner change rate. (**a**) No-cure scenario, (**b**) HIV remission scenario, and (**c**) HIV eradication scenario. The red circles correspond to the empirical cumulative histograms from the survey data. The blue lines correspond to the fitted Weibull distributions. The estimated shape and scale parameters of the distributions are listed in the legend of each panel. The vertical dashed lines indicate the intervals defining partner change rates for each risk group.

**Figure S3.**
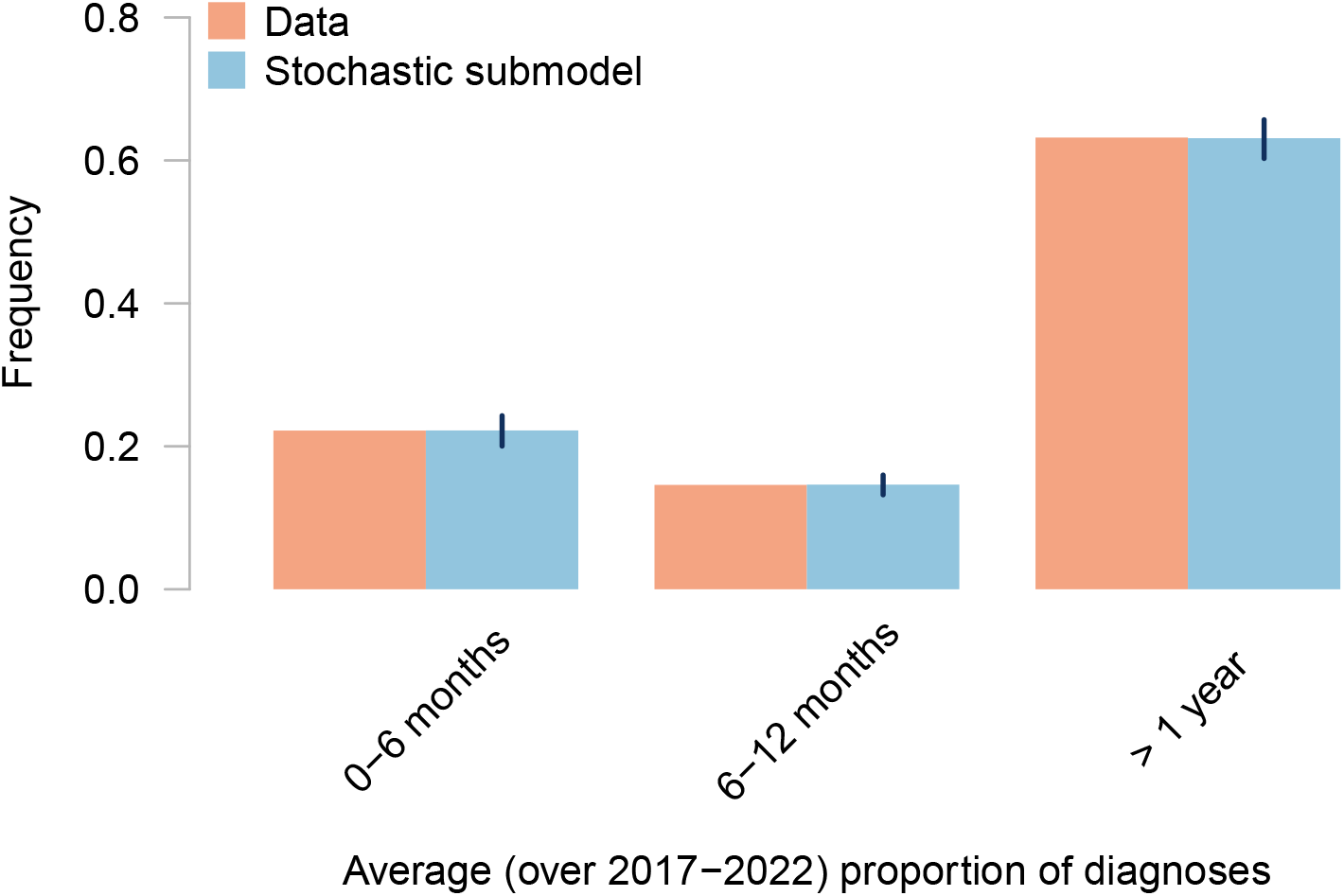
Model fitting against data on the proportion of diagnoses by time since HIV infection. The orange rectangles represent the SHM data. The blue rectangles and the error bars represent the mean and the 95% credible intervals for the proportions of diagnoses estimated from the stochastic model in Pseudocode 1 after calibration of the diagnosis rates.

**Figure S4.**
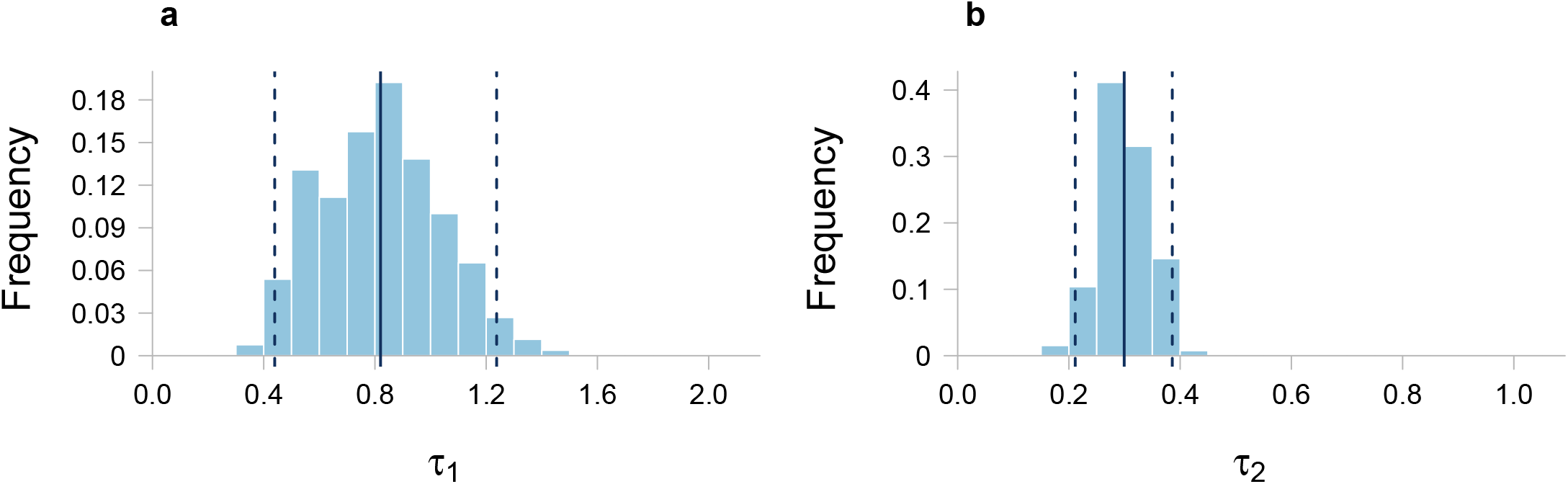
Estimated diagnosis rates. Histograms of diagnosis rates for (**a**) acute and (**b**) chronic HIV stages fitted via the Approximate Bayesian Computation framework. The vertical continuous line indicates the mean value. The vertical dashed lines indicate the bounds of the 95% credible interval.

**Figure S5.**
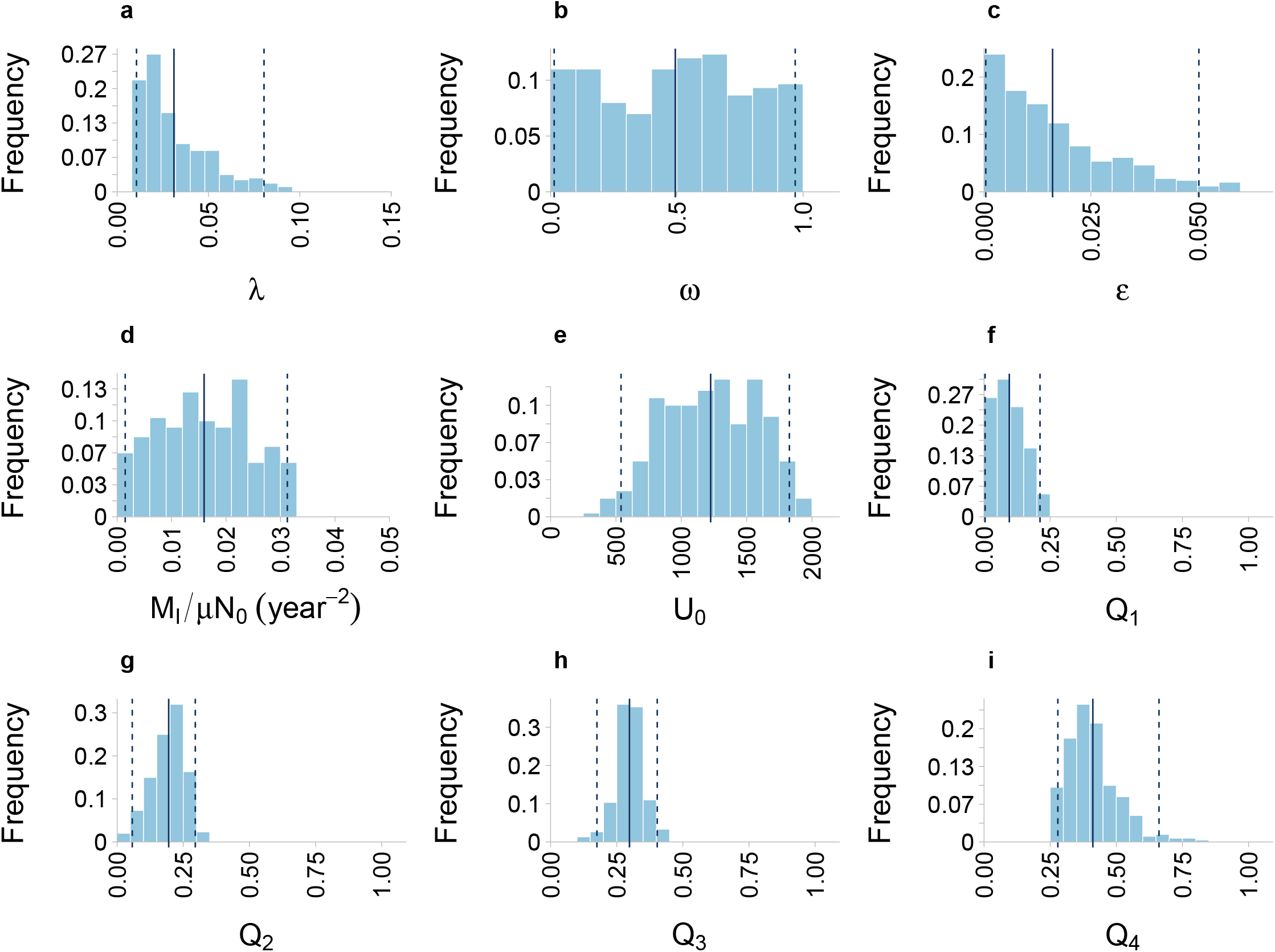
Estimated parameters. Histograms of all model parameters estimated via the Approximate Bayesian Computation framework. (**a**) Transmission probability per partner; (**b**) parameter describing mixing between risk groups; (**c**) relative infectivity of diagnosed and treated individuals; (**d**) number of newly imported undiagnosed HIV infections rescaled by the rate of entry into the sexually active population and the population size; (**e**) the number of undiagnosed individuals at the initialization of the model in 2015; (**f**) probability of imported infections being in the risk group 1; (**g**) probability of imported infections being in the risk group 2; (**h**) probability of the imported infections being in the risk group 3; (**i**) probability of the imported infections being in the risk group 4. The vertical continuous line indicates the mean value. The vertical dashed lines indicate the bounds of the 95% credible interval.

**Figure S6.**
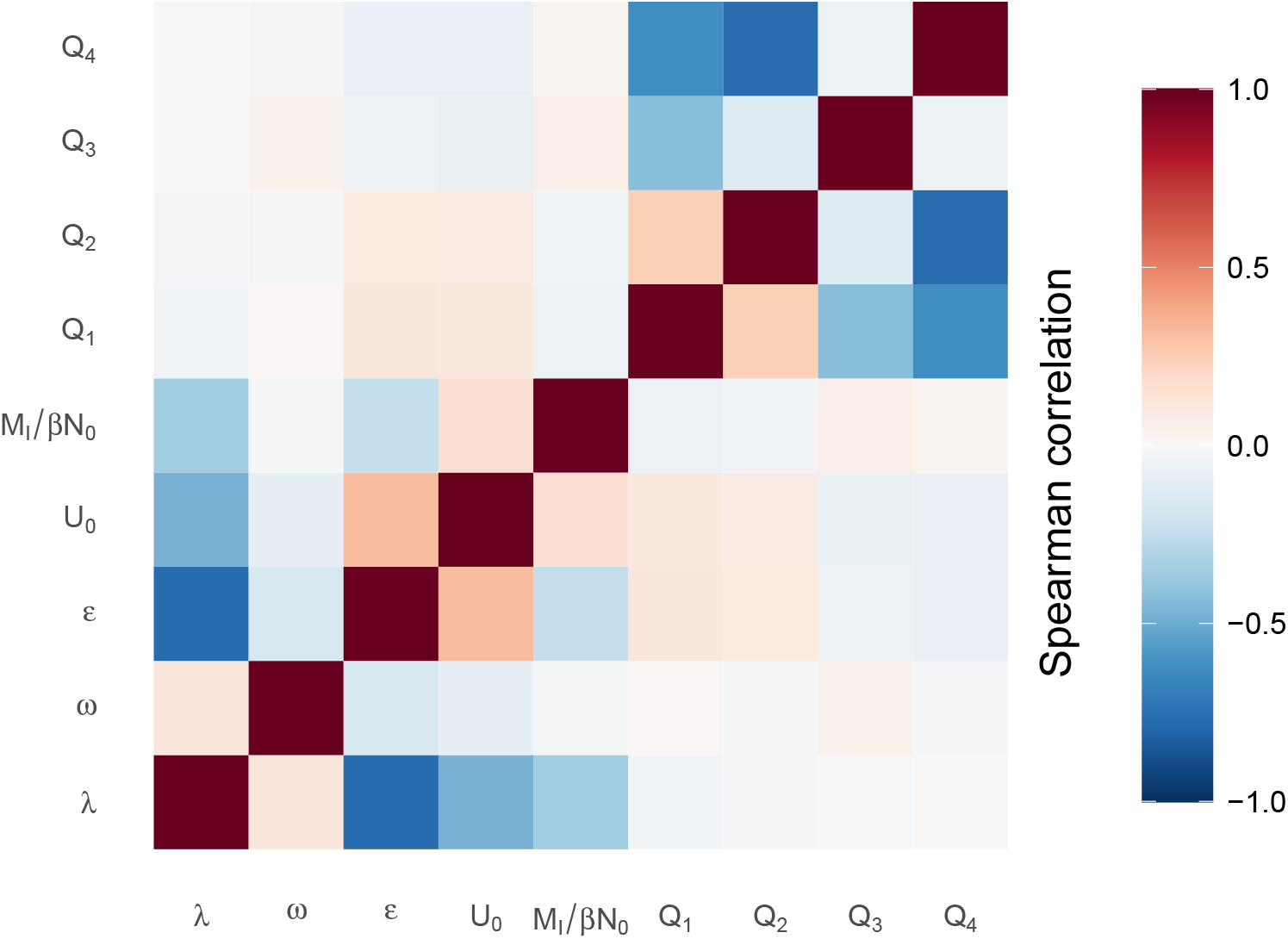
Correlation between estimated parameters. Spearman correlation for each pair of parameters estimated via the Approximate Bayesian Computation framework.

**Figure S7.**
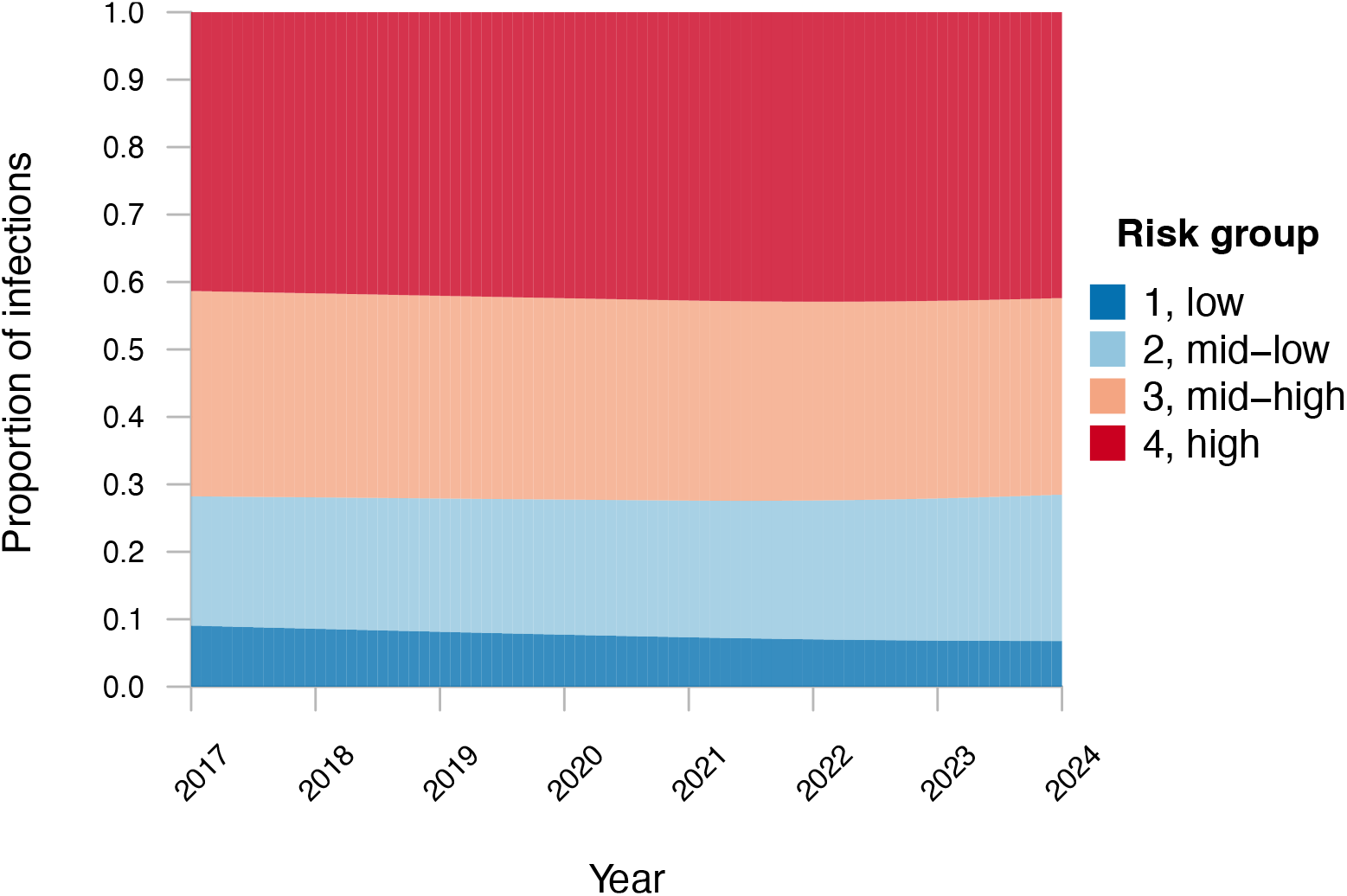
Proportions of infected individuals per risk group over time.

**Figure S8.**
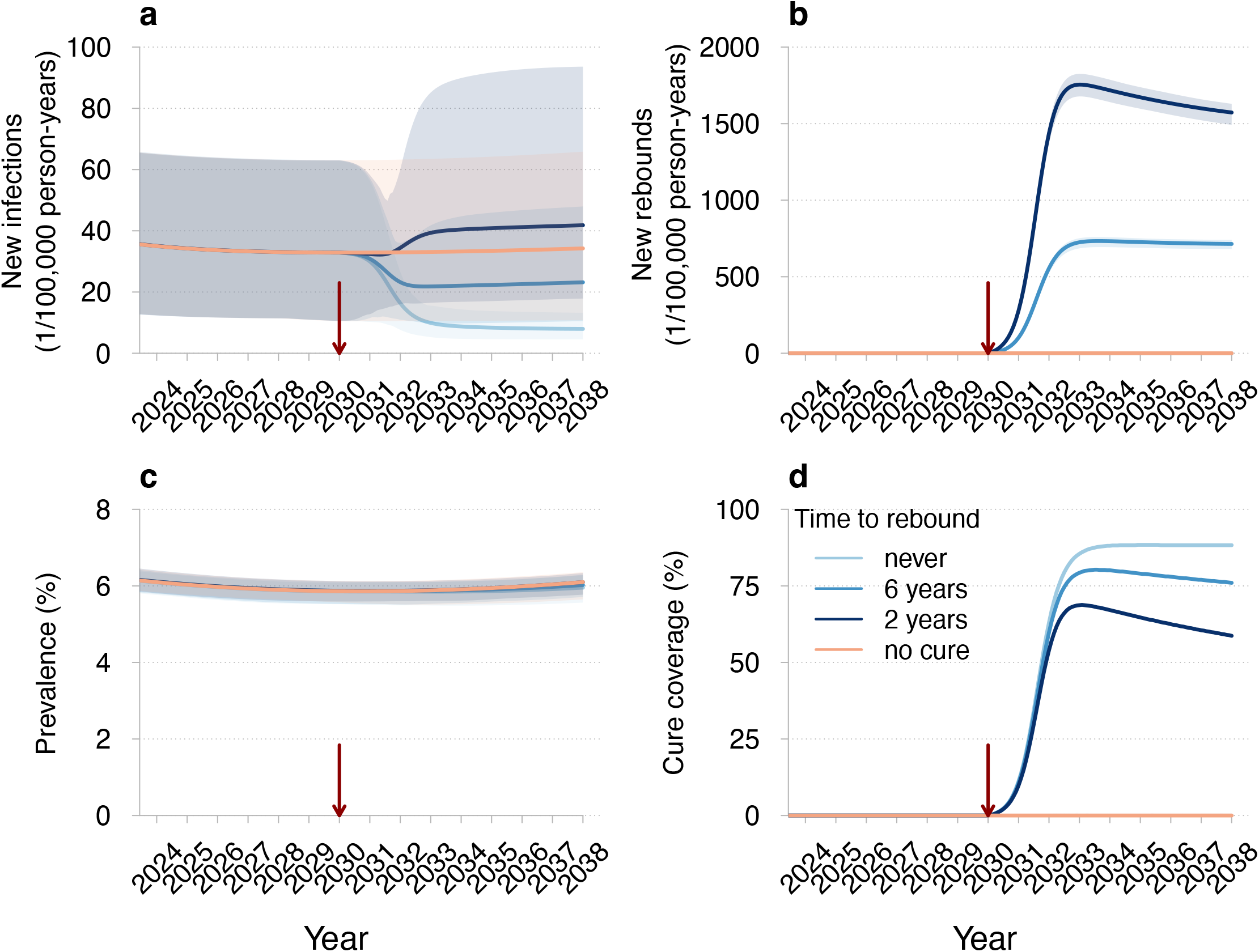
Projections of HIV dynamics under the HIV remission scenario if the intervention is introduced in 2030. This Figure can be compared with Figure 2 panel by panel. (**a**) New HIV infections, (**b**) new rebounds in individuals achieving HIV remission, (**c**) prevalence (proportion of individuals with HIV), and (**d**) cure coverage (proportion of eligible individuals achieving HIV remission) for different times until viral rebound. The red vertical arrows indicate the cure introduction. The mean trajectories from the model are shown as solid lines. The shaded regions correspond to 95% credible intervals based on 100 samples from the joint posterior parameter distribution. Different shades of blue correspond to different times until viral rebound. The projections of the model without a cure are shown in orange. The legend for different curves shown in (**d**) corresponds to all panels. Parameters: intervention efficacy of 90%, annual uptake of 90%, and diagnostic delay of individuals in HIV remission who experience a viral rebound of 3 months.

**Figure S9.**
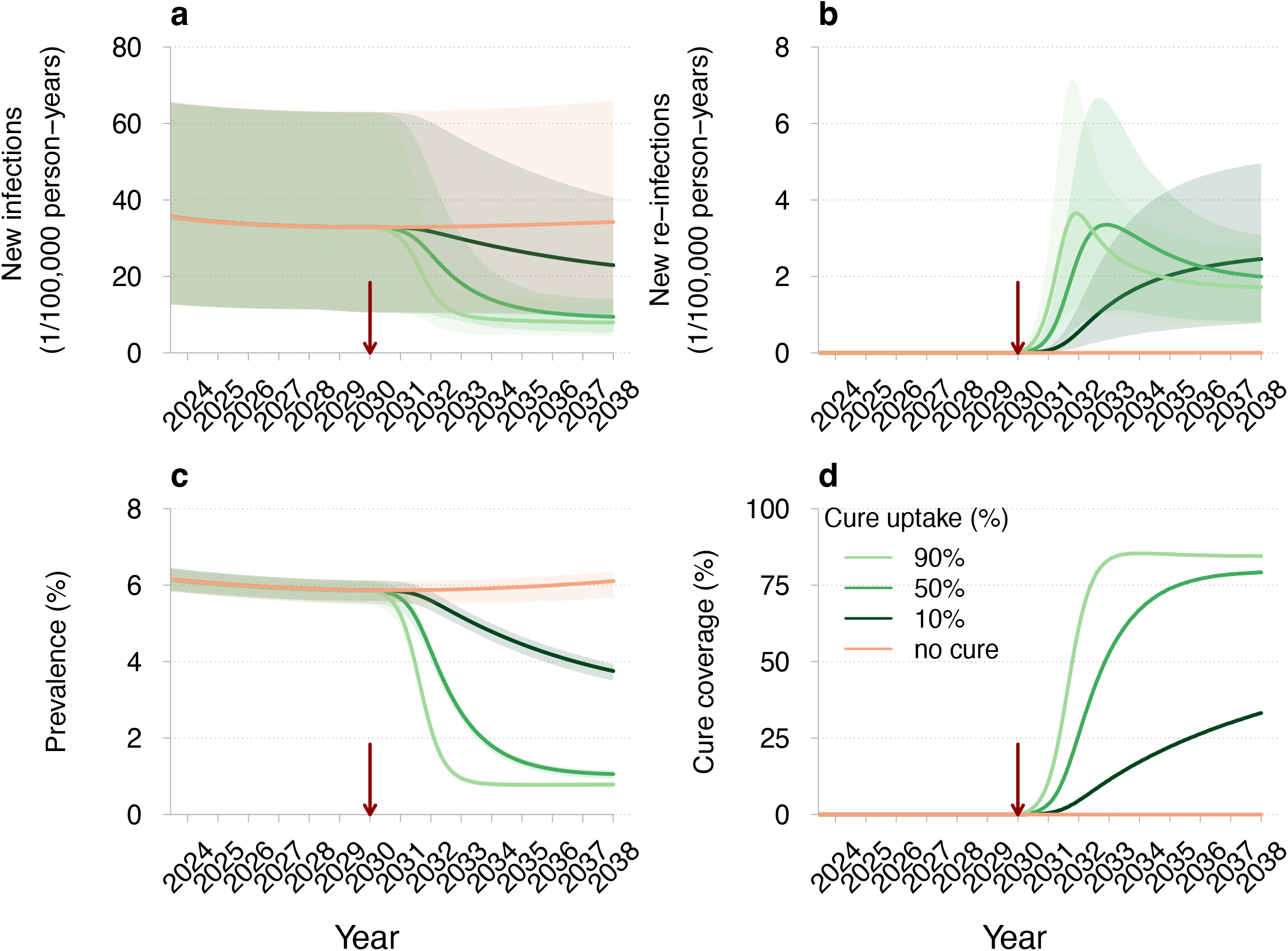
Projections of HIV dynamics under the HIV eradication scenario if the intervention is introduced in 2030. This Figure can be compared with Figure 4 panel by panel. (**a**) New HIV infections (primary infections in naive individuals), (**b**) new re-infections (secondary infections in cured individuals), (**c**) prevalence (proportion of individuals with HIV), and (**d**) cure coverage (proportion of cured individuals among all eligible) for different cure uptakes. The red vertical arrows indicate the cure introduction. The mean trajectories from the model are shown as solid lines. The shaded regions correspond to 95% credible intervals based on 100 samples from the joint posterior parameter distribution. Different shades of green correspond to different cure uptakes. The projections of the model without a cure are shown in orange. The legend for different curves shown in (**d**) corresponds to all panels. Parameters: cure efficacy of 90% and diagnostic delay of cured individuals who experience re-infection of 3 months.

**Figure S10.**
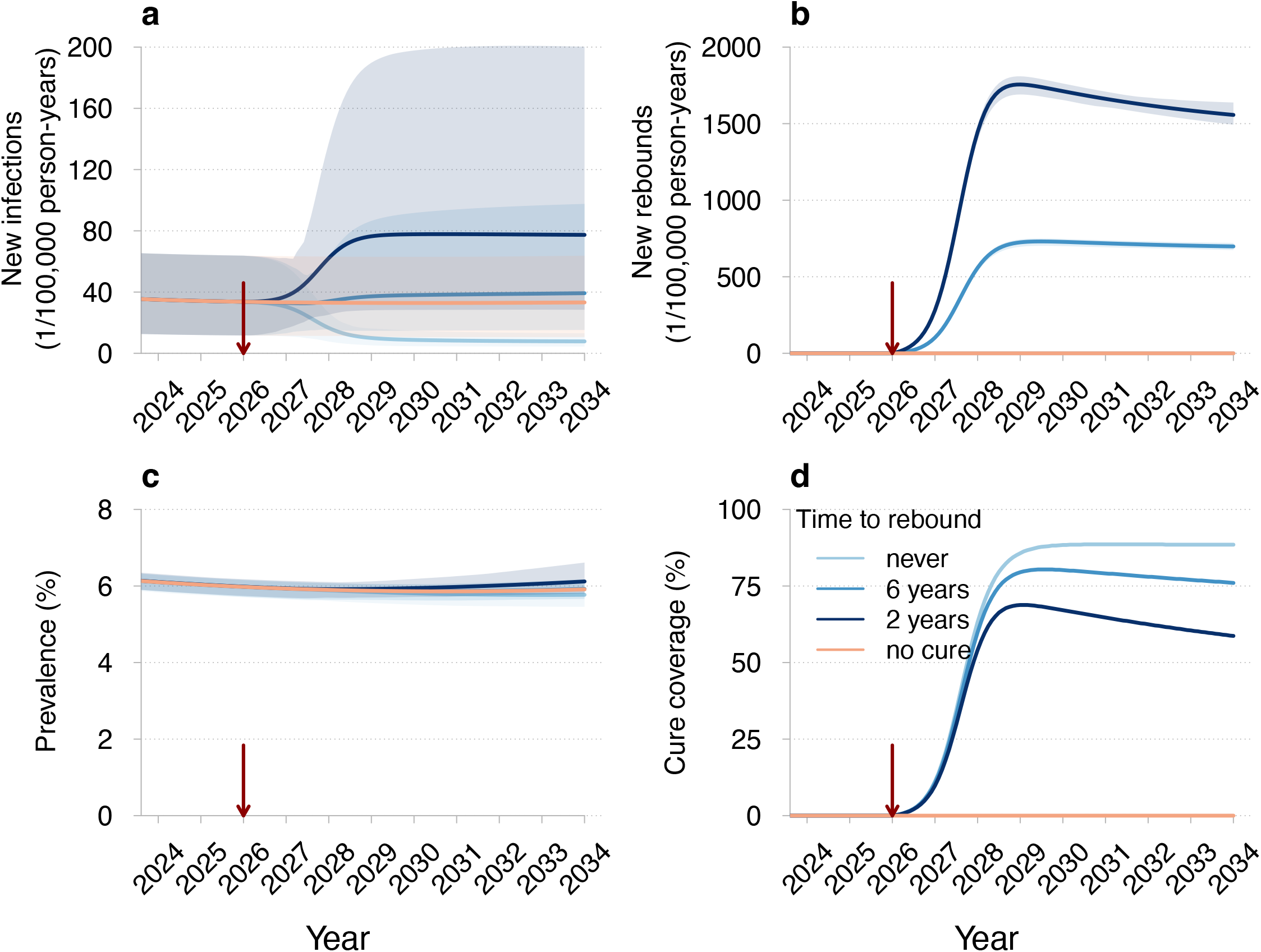
Projections of HIV dynamics under the HIV remission scenario if the first stage after a viral rebound is acute. (**a**) New HIV infections, (**b**) new rebounds in individuals achieving HIV remission, (**c**) prevalence (proportion of individuals with HIV), and (**d**) cure coverage (proportion of eligible individuals achieving HIV remission) for different times until viral rebound. The red vertical arrows indicate the cure introduction. The mean trajectories from the model are shown as solid lines. The shaded regions correspond to 95% credible intervals based on 100 samples from the joint posterior parameter distribution. Different shades of blue correspond to different times until viral rebound. The projections of the model without a cure are shown in orange. The legend for different curves shown in (**d**) corresponds to all panels. Parameters: intervention efficacy of 90%, annual uptake of 90%, and diagnostic delay of individuals in HIV remission who experience a viral rebound of 3 months.

**Figure S11.**
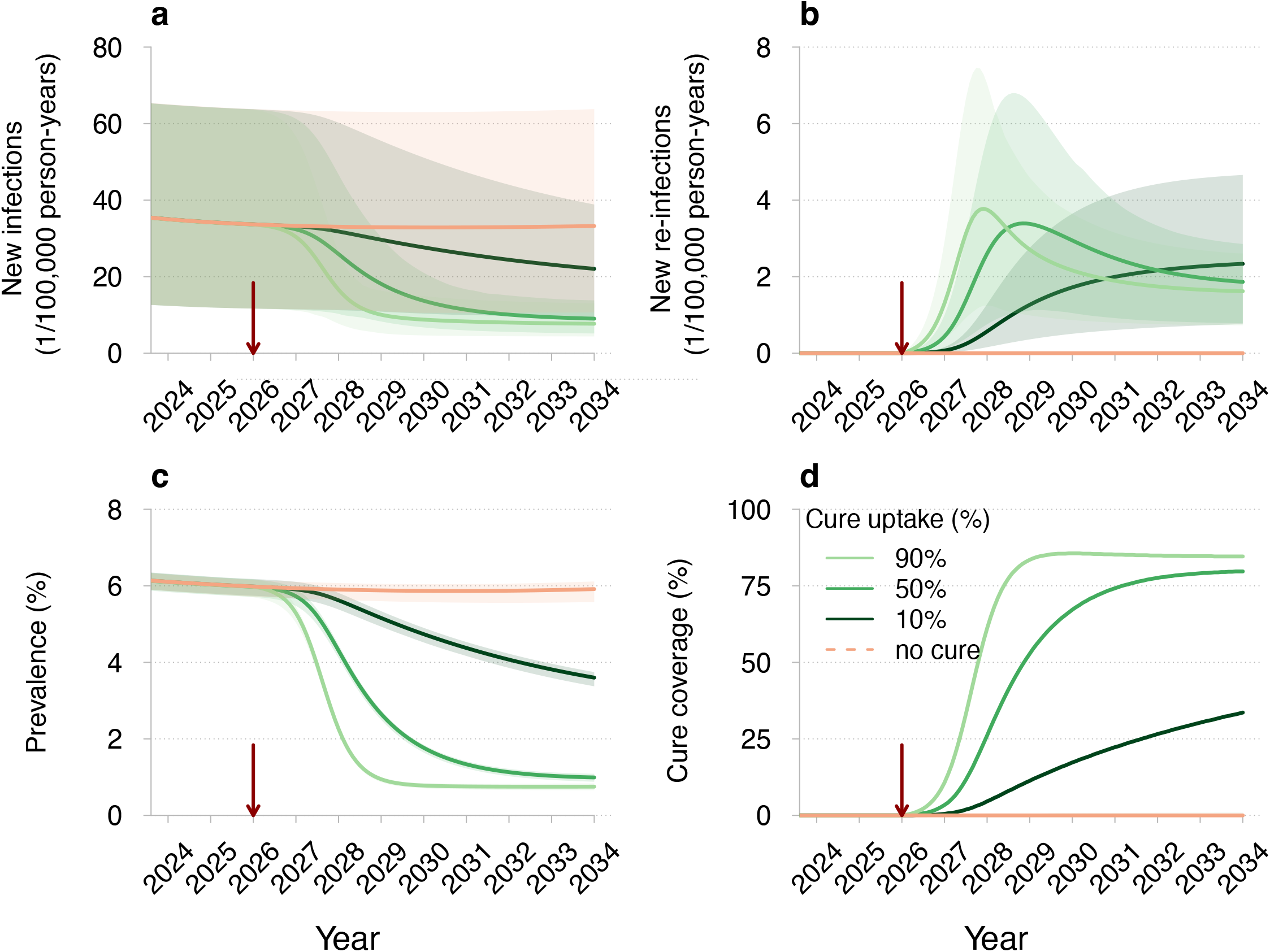
Projections of HIV dynamics under the HIV eradication scenario if the first stage after re-infection is chronic. This Figure can be compared with Figure 4 panel by panel. (**a**) New HIV infections (primary infections in naive individuals), (**b**) new re-infections (secondary infections in cured individuals), (**c**) prevalence (proportion of individuals with HIV), and (**d**) cure coverage (proportion of cured individuals among all eligible) for different cure uptakes. The red vertical arrows indicate the cure introduction. The mean trajectories from the model are shown as solid lines. The shaded regions correspond to 95% credible intervals based on 100 samples from the joint posterior parameter distribution. Different shades of green correspond to different cure uptakes. The projections of the model without a cure are shown in orange. The legend for different curves shown in (**d**) corresponds to all panels. Parameters: cure efficacy of 90% and diagnostic delay of cured individuals who experience re-infection of 3 months.

**Figure S12.**
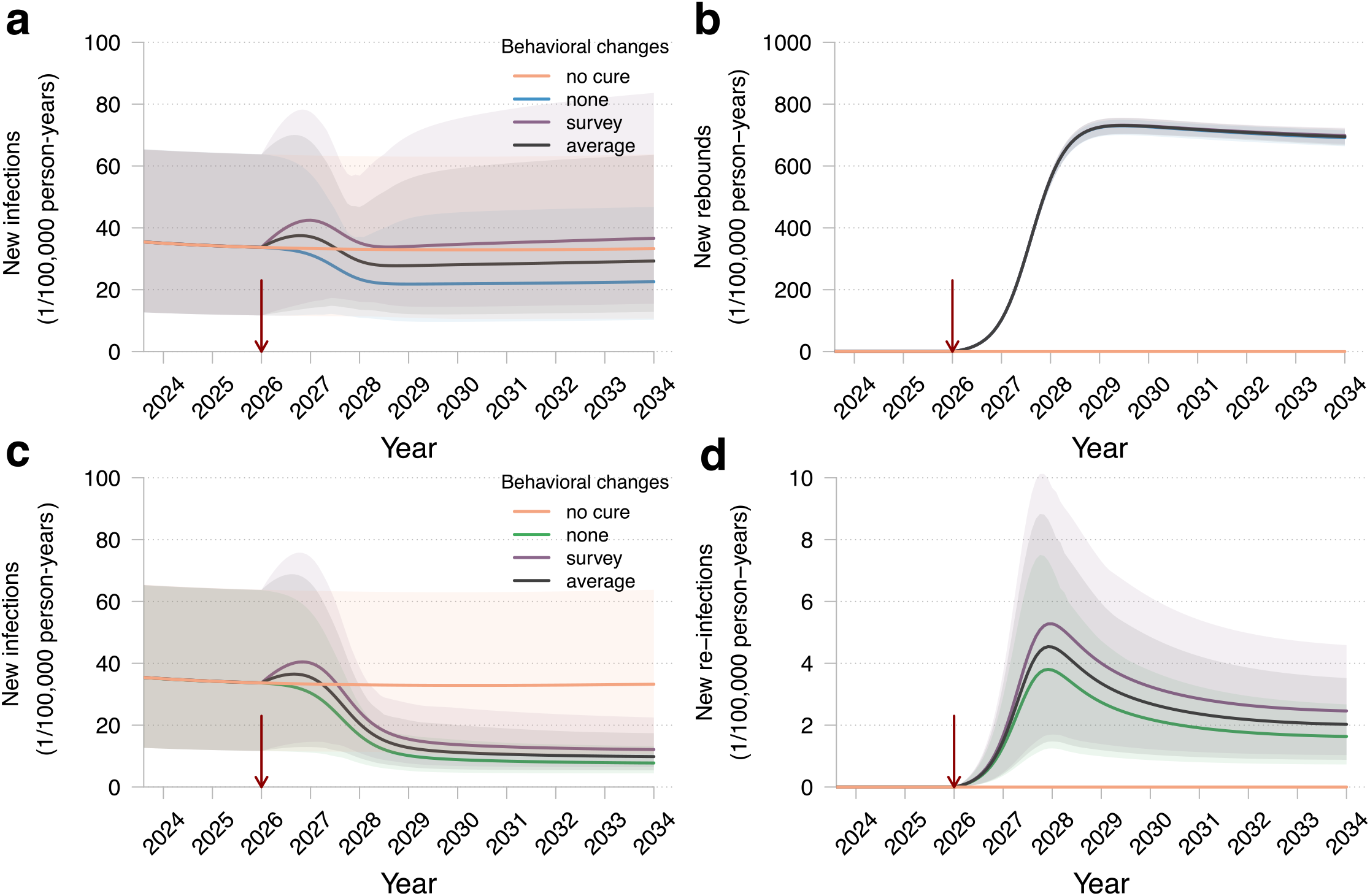
Projections of HIV dynamics under the HIV remission and HIV eradication scenarios assuming sexual behavior changes after cure. (**a,b**) HIV remission scenario with time to rebound of 6 years; (**c,d**) HIV eradication scenario. (**a,c**) New HIV infections (primary infections in naive individuals). (**b,d**) New rebounds/re-infections. Types of behavioral changes are shown in different colors: no changes (blue in (**a,b**) and green in (**c,d**)); survey partner change rates (purple), and average pre-cure and post-cure partner change rates (black). In (**d**) curves in different colors coincide. The red arrow indicates the cure introduction. The mean trajectories from the model are shown as solid lines. The shaded regions correspond to 95% credible intervals based on 100 samples from the joint posterior parameter distribution. The projections of the model without a cure are shown in orange. Parameters: cure efficacy of 90%, cure uptake of 90%, and diagnostic delay of cured individuals who experience a viral rebound or re-infection of 3 months.

**Figure S13.**
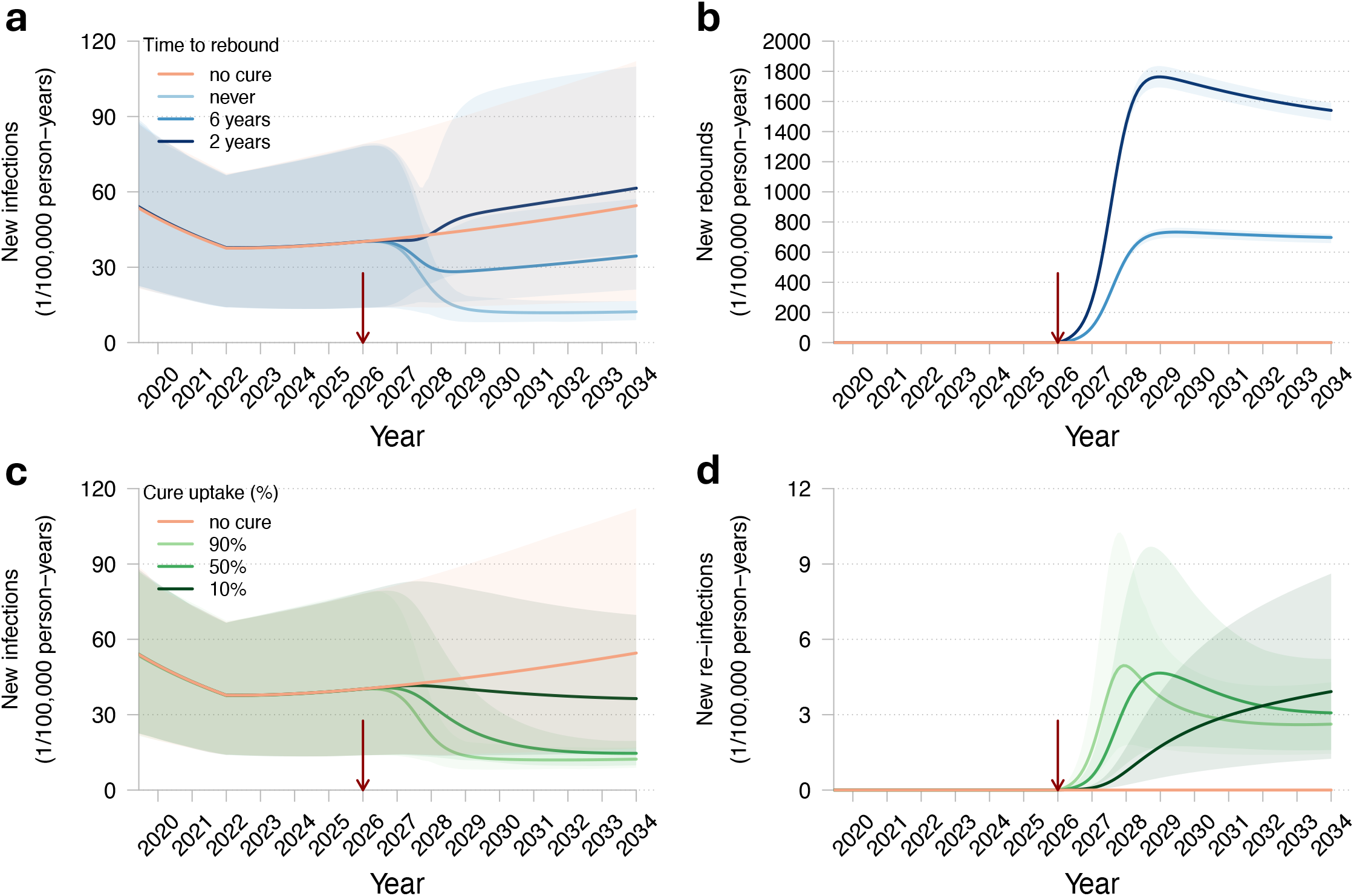
Projections of HIV dynamics under the HIV remission and HIV eradication scenarios assuming an increase in HIV incidence starting in 2022. (**a,b**) HIV remission scenario. Different shades of blue correspond to different times until viral rebound. Cure uptake is set at 90% in the HIV remission scenario. (**c,d**) HIV eradication scenario. Different shades of green correspond to different cure uptakes. (**a,c**) New HIV infections (primary infections in naive individuals). (**b,d**) New rebounds/re-infections. The red arrow indicates the cure introduction. The mean trajectories from the model are shown as solid lines. The shaded regions correspond to 95% credible intervals based on 100 samples from the joint posterior parameter distribution. The projections of the model without a cure are shown in orange. Parameters common to all panels: cure efficacy of 90%, and diagnostic delay of cured individuals who experience a viral rebound or re-infection of 3 months.

